# Factors associated with higher levels of grief and support needs among people bereaved during the pandemic: Results from a national online survey

**DOI:** 10.1101/2022.02.07.22270593

**Authors:** LE Selman, D J J Farnell, M Longo, S Goss, A Torrens-Burton, K Seddon, C R Mayland, L Machin, A Byrne, E J Harrop

**Affiliations:** Cardiff University, School of Dentistry, Cardiff, UK; Cardiff University, Marie Curie Research Centre, Cardiff, UK; Cardiff University, PRIME Centre, Division of Population Medicine, Cardiff, UK; Wales Cancer Research Centre, Cardiff, UK; University of Sheffield, Department of Oncology and Metabolism, Sheffield, UK; Keele University, Keele, UK

**Author notes:** Correspondence to: Lucy Selman, University of Bristol, Palliative and End of Life Care Research Group, Population Health Sciences, Bristol Medical School, Canynge Hall, 39 Whatley Road, Bristol BS8 2PS, U.K.

## Abstract

**Background:** The COVID-19 pandemic has affected millions of people’s experiences of bereavement. We aimed to identify risk factors for grief and support needs.

**Methods:** Online survey of people bereaved in the UK (deaths 16 March 2020-2 January 2021), recruited via media, social media, national associations/organisations. Grief was assessed using the Adult Attitude to Grief (AAG) scale, which calculates an overall index of vulnerability (IOV) (range 0-36). Practical and emotional support needs were assessed in 13 domains.

**Results:** 711 participants, mean age 49.5 (SD 12.9, range 18-90). 628 (88.6%) were female. Mean age of the deceased 72.2 (SD 16.1). 311 (43.8%) deaths were from confirmed/suspected COVID-19. Mean IOV was 20.41 (95% CI = 20.06 to 20.77). 28.2% exhibited severe vulnerability (IOV ≥ 24). In six support domains relating to psycho-emotional support, 50% to 60% of respondents reported high/fairly high levels of need. Grief and support needs increased strongly for close relationships with the deceased (versus more distant) and with reported social isolation and loneliness (*P* < 0.001), whereas they reduced with age of the deceased above 40 to 50. Other risk factors were place of death and reduced support from health professionals after death (*P* < 0.05).

**Conclusions:** High overall levels of vulnerability in grief and support needs were observed. Relationship with the deceased, age of the deceased, and social isolation and loneliness are potential indicators of those at risk of even higher vulnerability in grief and support needs. Healthcare professional support after death is associated with more positive bereavement outcomes.

## Introduction

Over 5.7 million people have died of COVID-19 globally (as of February 2022), leaving over 51 million people bereaved (Verdery *et al*., 2020). The COVID-19 pandemic has had a detrimental impact on end-of-life and bereavement experiences, with infection control and social distancing restrictions impacting usual processes of saying goodbye and mourning a death, regardless of the cause of death. Lack of access to, and physical contact with, family and friends at the time of death, and restrictions surrounding funerals have caused high levels of distress to those bereaved during the pandemic (Hanna *et al*., 2021, Torrens-Burton *et al*., 2021, Yardley and Rolph, 2020). Studies during (Breen *et al*., 2021a, Eisma *et al*., 2021, Mayland *et al*., 2021) and before the pandemic (Harrop *et al*., 2016, Kentish-Barnes *et al*., 2015, Kristensen *et al*., 2012) have found that traumatic end-of-life experiences and sudden deaths, which are common in COVID-19, exacerbate the severity of grief experiences. Societal disruption and limited access to usual support networks also increase risks of poor bereavement outcomes (Lobb *et al*., 2010, Smith *et al*., 2020), although the impact of the pandemic in this regard is not yet fully understood.

To inform clinicians and bereavement support providers and direct resource allocation, evidence is needed to identify groups at particular risk of difficulties in their grief and high levels of support needs. To date, three relevant studies have been published, one in China (Tang and Xiang, 2021, Tang *et al*., 2021) and two in the USA (Breen *et al*., 2021a, Breen *et al*., 2021b, Neimeyer and Lee, 2021). The first study, in China, examined risk factors for prolonged grief disorder among people bereaved by COVID-19, but did not examine the impact of pandemic-related circumstances such as social isolation. The US studies examined associations between pandemic-related complications, demographic characteristics, functional impairment and dysfunctional grief symptoms. However, findings across these initial studies are inconsistent (e.g. regarding the impact of COVID-19 deaths on bereavement), and none have examined risk factors for participants’ self-reported support needs.

We conducted a longitudinal mixed-methods study of bereavement during the pandemic in the UK. In previous papers, we described sub-optimal end-of-life care, challenging experiences after bereavement, support needs and barriers to accessing support among 711 people bereaved of any cause (Harrop *et al*., 2021, Selman *et al*., 2021b, Torrens-Burton *et al*., 2021). In this paper, we aim to identify factors associated with higher levels of grief and bereavement support needs, using cross-sectional baseline data. Our research question was: which pandemic-related challenges and demographic and clinical characteristics are associated with higher levels of grief and support needs among people in the UK bereaved during the COVID-19 pandemic?

## Methods

### Study design

An open web survey (Supplementary file 1) was disseminated to a convenience sample of people bereaved during the pandemic in the UK.

### Survey development

Survey items and structure were informed by study aims and previous research (Claessen *et al*., 2013, Harrop *et al*., 2020a, Harrop *et al*., 2020b, Sue Ryder, 2019). The survey was designed with input from a multi-professional advisory group and piloted, refined and tested with 16 members of the public with experience of bereavement. Non-randomised open and closed questions covered end-of-life and grief experiences, and perceived needs for, access to, and experiences of bereavement support.

### Primary outcomes

*Grief* was assessed using the 9-item Adult Attitude to Grief (AAG) scale (Sim *et al*., 2014). The scale is based on the Range of Response to Loss model (Machin, 2001, 2013), which identifies three distinct responses: being ‘overwhelmed’, a state dominated by emotional and cognitive distress; being ‘controlled’, a state dominated by the need to avoid emotional expression and focus on day-to-day life; and being balanced or ‘resilient’, expressed through feeling supported and able to cope. AAG subscale scores indicate levels of feeling overwhelmed, controlled, and reversed resilience on a scale of 0 (none) to 12 (very high). An overall index of vulnerability (IOV) is calculated by adding these subscale scores together (IOV: 0-20 = low vulnerability, 21-23 = high vulnerability, and 24-36 = severe vulnerability (Sim *et al*., 2014)). This questionnaire has previously been validated and psychometrically tested (Sim *et al*., 2014). We also found acceptable levels of internal consistency/reliability (Cronbach’s α = 0.73, 0.70, and 0.76, respectively).

*Support needs* were assessed in 13 domains, informed by previous studies (Harrop *et al*., 2020a, Harrop *et al*., 2020b) (Table S3). Two subscales (emotional support and practical support) were found via exploratory factor analysis. Cronbach’s α was given by 0.79, 0.95, and 0.94 for the practical support subscale, emotional support subscale, and all items, respectively, indicating high levels of reliability/internal consistency. Subscale scores are found by determining the mean across all items in a given subscale. The overall mean is evaluated over all 13 items. We interpret results for both subscale scores and the overall mean score via: 1 = no support needed; 3 = moderate level of support needed; 5 = high level of support needed.

### Associated factors

We assessed whether demographic and clinical factors, experiences of end-of-life care and pandemic-related problems independently predicted levels of grief and support needs. Factors included in the analysis are recognised risk factors for poor bereavement outcomes (age, gender, relationship to deceased, expectedness of the death, ability to say goodbye to the deceased, experiences of end-of-life care, perceived social support) (Kentish-Barnes *et al*., 2015, Lobb *et al*., 2010, Selman *et al*., 2020, Yamaguchi *et al*., 2017) or have been identified to be indirectly associated with such outcomes (qualifications, deprivation level and region; place of death; cause of death) (Haugen *et al*., 2021, Miyashita *et al*., 2008). We used postcode data to identify geographical region of residence and (for England) socio-economic deprivation.

#### Experiences of end-of-life care

Six items, adapted from the Consumer Quality Index for Palliative Care (Claessen *et al*., 2013), assessed end-of-life care experiences: involvement in care decisions, knowing the contact details for the professional responsible for care, receiving information about the approaching death, support by healthcare professionals immediately after the death, contact by the hospital/care provider after the death, and provision of information about bereavement support services.

#### Pandemic-related problem

Six items assessed pandemic-related challenges prior to and after the death, e.g. being unable to visit the person who died prior to their death, restricted funeral arrangements, social isolation and loneliness (see Table S8). All items were answered yes/no. Respondents were asked to tick all the experiences that applied to them.

### Study procedure

The survey was administered via Jisc software (JISC, 2021), open 28th August 2020 to 5th January 2021 and disseminated on social and mainstream media and via voluntary sector associations and bereavement support organisations, including organisations representing ethnic minority communities. Organisations helped disseminate the voluntary survey by sharing on social media, web-pages, newsletters, on-line forums and via direct invitations to potential participants (see example advertisement, Supplementary file 2). For ease of access, the survey was posted onto a bespoke study-specific website with a memorable URL (covidbereavement.com). Two participants chose to complete the survey in paper format. Summaries of survey results (including interim results released November 2020 (Harrop E., 2020)) were posted on the website.

Inclusion criteria: aged 18+; family or close friend bereaved since social-distancing requirements were introduced in the UK (16/03/2020); death occurred in the UK; ability to consent. The initial section of the survey requested informed consent and provided data protection information (see Supplementary file 1). 12 surveys were completed in duplicate; the first completed survey was retained for these participants. Two incomplete surveys were excluded where only the consent question had been answered.

Reporting follows the Checklist for Reporting Results of Internet E-Surveys (Eysenbach, 2004).

### Data analysis

A statistical analysis plan was drafted by the study statistician (DJJF) and refined iteratively by all members of the research team. Descriptive statistics were used to describe all variables, with normality of outcomes assessed as appropriate. Standard univariate statistical tests were used first to compare differences between groups for all outcomes. A standardised effect size (Cohen’s *d*) was used to measure differences between groups for ordinal or continuous variables (*d* = 0.3: small effect, *d* = 0.5: medium effect, *d* = 0.8: large effect, *d* = 1.2: very large effect). Where factors contained more than one group, we used the maximum difference in means between any two groups in the factor and the average standard deviation across all groups. By using a standardised measure of effect size, the effects of factor on outcomes could be compared directly and patterns across multiple outcomes ascertained. Factors with consistently medium or large effects across multiple outcomes were included in a mixed model of IOV, which complements results of univariate analyses and adjusts for any effects of confounding. A directed acyclic graph (DAG) was also used to map out the relationships between variables, which was useful in planning the statistical analyses and visualising the mixed-effects model. No readily apparent pattern occurred in IOV with respect to region of the UK and so this was taken to be the random effect, whereas all other factors were taken as fixed effects. IOV reduced with age of the deceased above 40 to 50 years old and so this was introduced into the mixed model as an explicit (quadratic) covariate. Interactions between variables in the model did not improve the model fit significantly or change regression coefficients greatly. Residuals for this model were normally distributed, as required, and all assumptions of this approach were satisfied. All calculations were carried out using SPSS V26.

### Ethical approval

The study was approved by Cardiff University School of Medicine Research Ethics Committee (SMREC 20/59) and conducted in accordance with the Declaration of Helsinki and all respondents provided informed consent.

## Results

### Sample characteristics

711 bereaved participants completed the survey (Table 1). Participants represented diverse geographical areas, deprivation indexes and levels of education. 88.6% of participants were female (*n* = 628); the mean age of the bereaved person was 49.5 years old (SD = 12.9; range 18-90). The most common relationship of the deceased to the bereaved was parent (*n* = 395, 55.6%), followed by partner/spouse (*n* = 152, 21.4%). 72 people (10.1%) had experienced more than one bereavement. 33 people (4.7%) self-identified as from a minority ethnic background. Missing data was minimal (i.e., close to zero) for all variables and so imputation was not necessary.

**Table 1:**
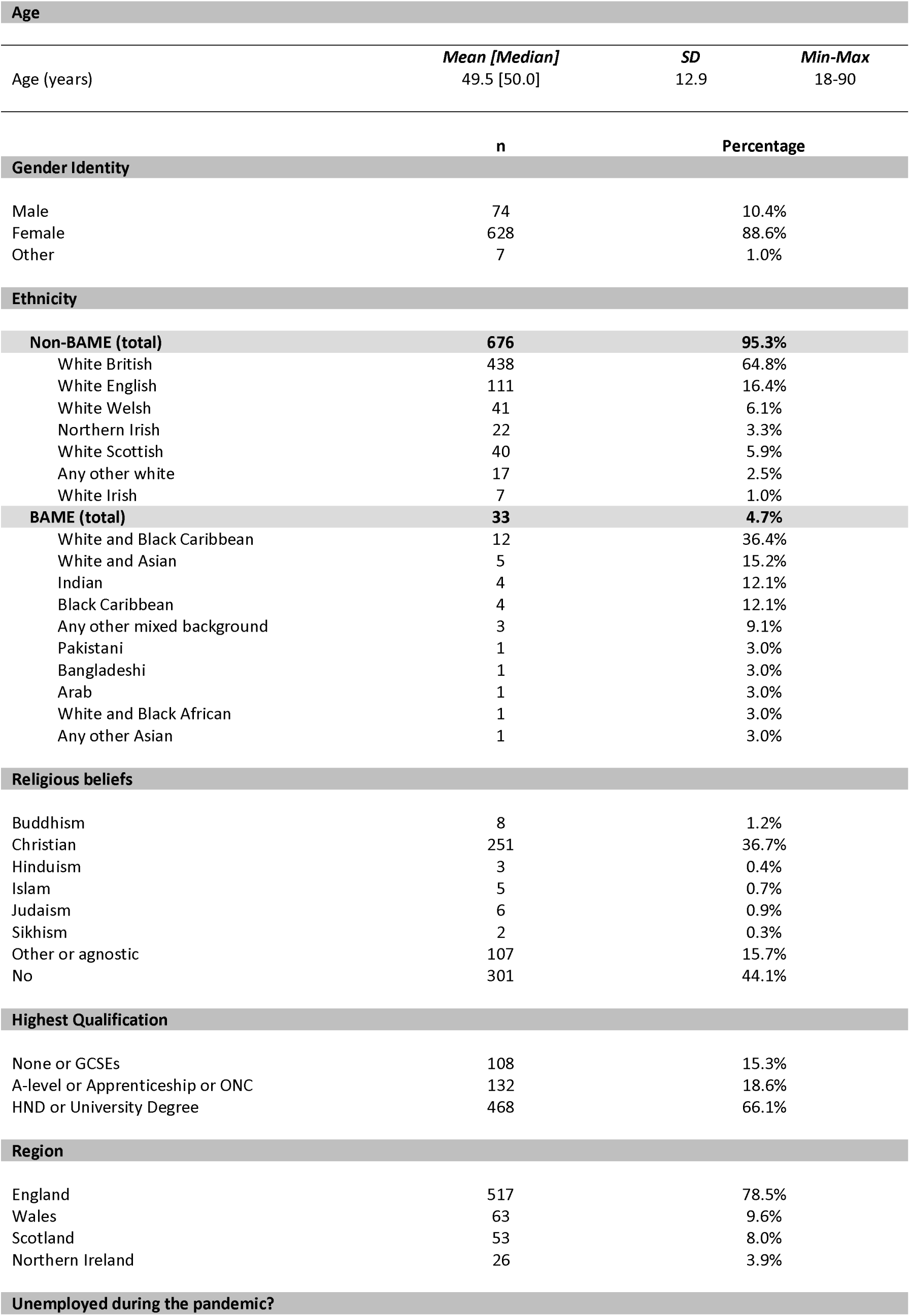
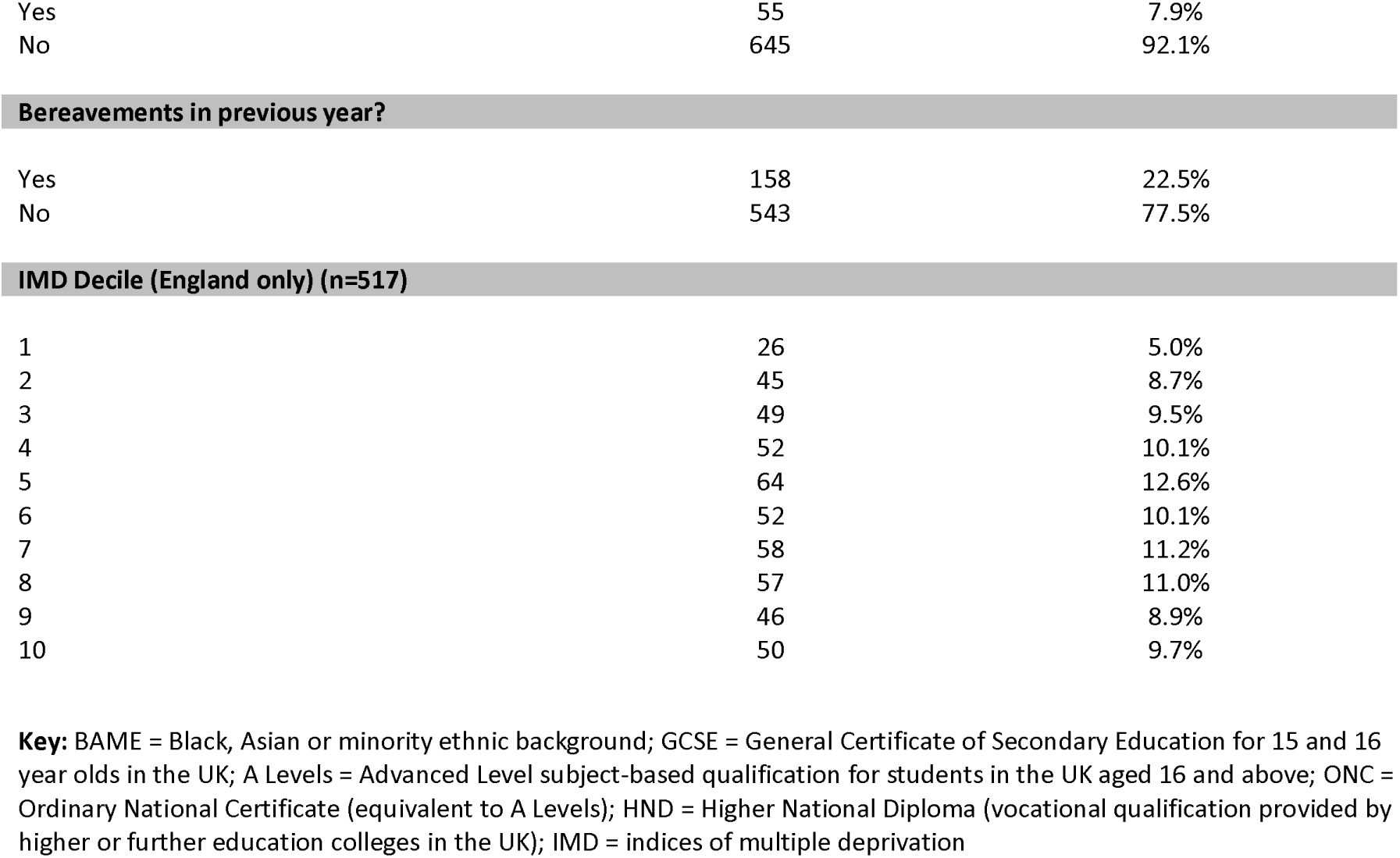
Characteristics of the bereaved person.

Table 2 presents the characteristics of the deceased people. The mean age of the deceased person was 72.2 years old (SD=16.1; range 4 months gestation to 102 years). 43.8% (*n* = 311) died of confirmed or suspected COVID-19, 21.9% (*n* = 156) from cancer, and 16.7% (*n* = 119) from another life-limiting condition. Most died in hospital (*n* = 410, 57.8%). Questionnaires were completed a median of 152 days (5 months) after the death (range 1-279 days).

**Table 2:** Characteristics of the deceased.

| Age |  |  |  |
| --- | --- | --- | --- |
|  | *Mean [Median]* | *SD* | *Min-Max* |
| Age (years) | 72.2 [74.0] | 16.1 | 0-102 |

|  | **n** | **Percentage** |
| --- | --- | --- |
| Relationship of the Deceased Person to the Bereaved* |  |  |
| Partner | 152 | 21.4% |
| *(Male / Female)* | *(129 / 23)* | *(18.1% / 3.2%)* |
| Parent | 395 | 55.6% |
| *(Father / Mother)* | *(218 / 197)* | *(30.7% / 27.7%)* |
| Grandparent | 54 | 7.6% |
| Sibling | 23 | 3.2% |
| *(Brother / Sister)* | *(15 / 10)* | *(2.1%, 1.4%)* |
| Child | 15 | 2.1% |
| *(Son / Daughter)* | *(12 / 4)* | *(1.7% / 0.6%)* |
| Other family member | 46 | 6.5% |
| Colleague or friend | 26 | 3.7% |

| Cause of Death |  |  |
| --- | --- | --- |
| COVID | 273 | 38.5% |
| Suspected COVID | 38 | 5.4% |
| Non-COVID (total) | 399 | 56.2% |
| *Cancer* | 156 | 21.9% |
| *Other PLLC\*\** | 118 | 16.7% |
| *Non-PLLC/SD\*\*\** | 112 | 15.8% |
| *Don't know* | 12 | 3.0% |
| *Not specified* | 1 | 0.2% |

| Was the death expected? |  |  |
| --- | --- | --- |
| Yes | 113 | 16.0% |
| No | 552 | 78.0% |
| Don't know | 43 | 6.1% |

| Place of death |  |  |
| --- | --- | --- |
| In hospital | 410 | 57.7% |
| In their home | 158 | 22.2% |
| In a hospice | 37 | 5.2% |
| In a care home | 91 | 12.8% |
| Other/Don't know | 13 | 1.8% |

\* Multiple bereavements recorded by participants explain discrepancies between overall totals in sibling, child and parent groups and their sub-categories.
\*\*PLLC = Progressive life-limiting condition e.g. heart disease, COPD, dementia
\*\*\*Non-PLLC/SD = Non-progressive life-limiting condition/sudden death e.g. stroke, heart attack, accident, suicide

### Primary outcomes

#### Levels of grief

Individual AAG item and subscale scores are given in Supplementary Tables S1-S2. Mean IOV was 20.41 (95% CI = 20.06 to 20.77, median = 21.00), i.e., demonstrating high levels of vulnerability in grief overall. 48.4% exhibited low levels of vulnerability (i.e., 0 ≤ IOV ≤ 20); 23.4% exhibited high levels of vulnerability (i.e., 21 ≤ IOV ≤ 23), and 28.2% severe levels of vulnerability (i.e., IOV ≥ 24). Overall subscale scores were: overwhelmed mean = 8.53 (95% CI = 8.31 to 8.72), controlled mean = 6.61 (95% CI = 6.41 to 6.82), reversed resilience mean = 5.28 (95% CI = 5.07 to 5.49).

#### Support needs

In 6 (of 13) domains, all relating to psycho-emotional support, between 50% to 60% of respondents reported high/fairly high levels of need (Table S3). The three most common of these were: dealing with my feelings about the way my loved one died (60%), expressing my feelings and feeling understood by others (53%), and feelings of anxiety and depression (53%). Subscale scores were: emotional subscale mean = 3.33 (95% CI = 3.25 to 3.41), i.e., moderate level of emotional support needed; practical subscale, mean = 2.41 (95% CI = 2.34 to 2.50), i.e., low to moderate level of practical support needed. Overall support (over all items) demonstrated a mean = 3.12 (95% CI = 3.04 to 3.19), i.e., a moderate level of support needed overall.

#### Factors associated with levels of grief and support needs

Effect sizes measured via Cohen’s *d* were used to determine those factors and covariates that had the strongest influence on the outcome measures. We describe significant effects of these factors on the outcomes in detail below, by magnitude of effect (also see Tables 3–4 and Supplementary Tables S4-S10).

**Table 3:**
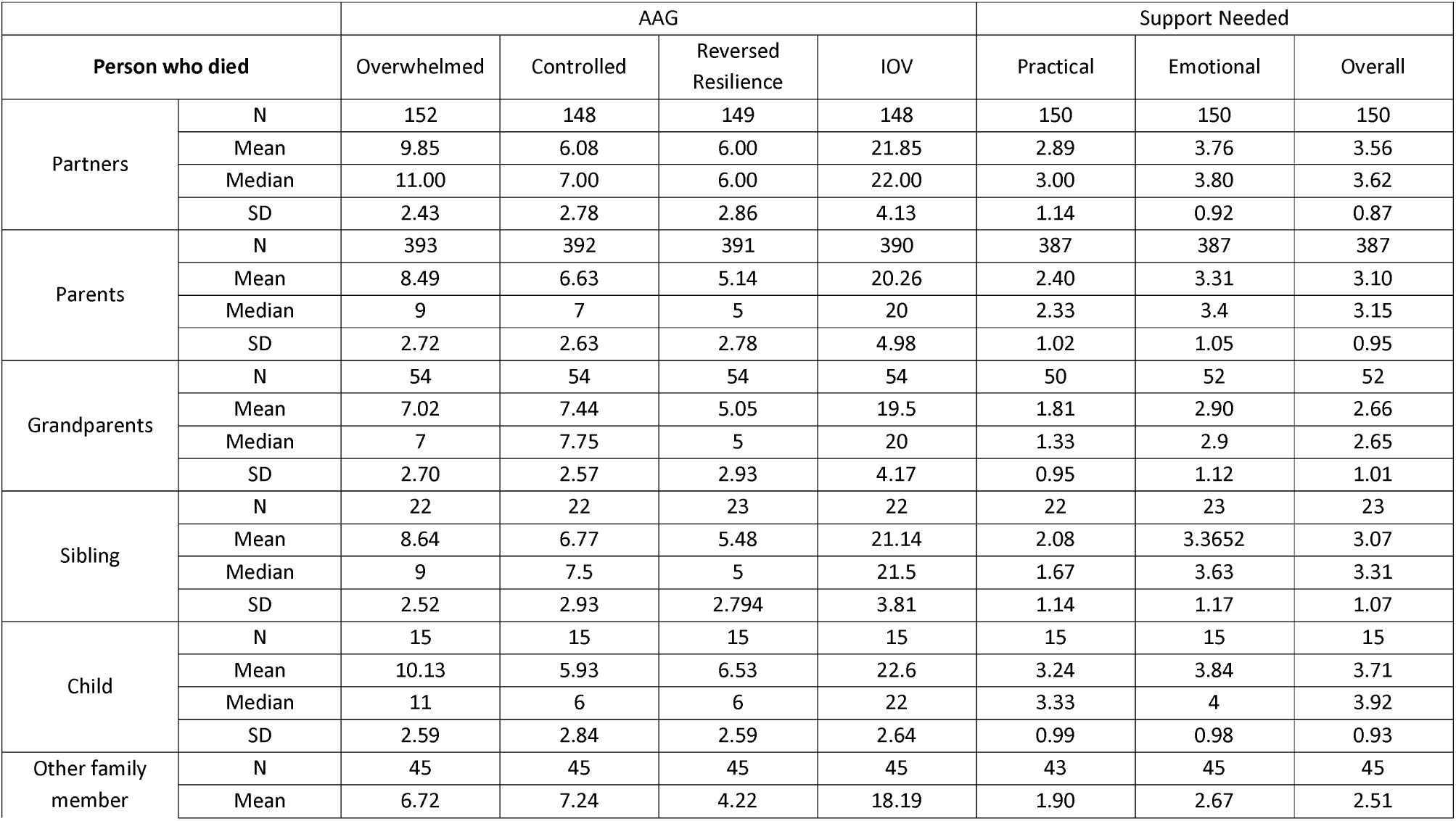
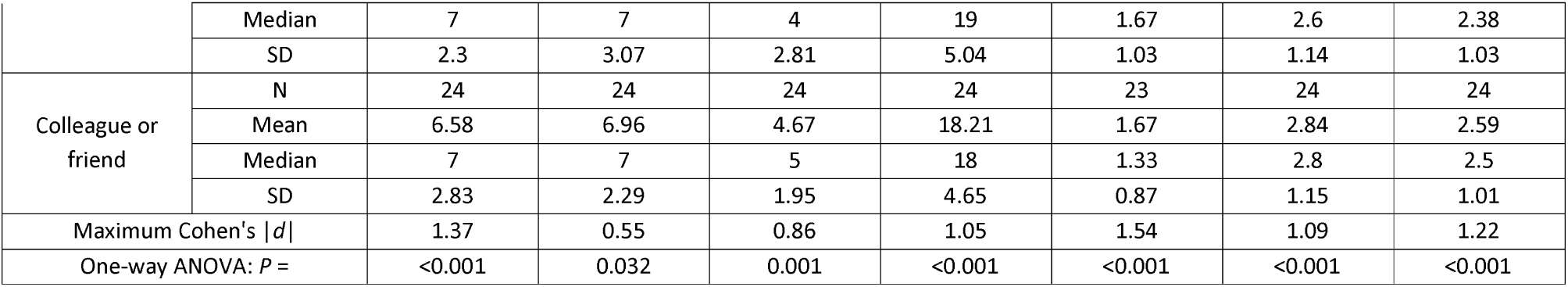
Results for subscales and scale scores for the AAG questionnaire and support needed as a function of the relationship between the bereaved person and the deceased.

##### Relationship between the bereaved and the deceased

Strong differences (*P* < 0.001) occur for all AAG subscales and IOV and practical, emotional, and overall support needs as a function of the relationship of the deceased person to the bereaved. In particular, IOV, emotional and overall support need scores were much higher for close family, particularly when the person who died was a child or partner (followed by sibling or parent), compared with more distant family members and colleagues or friends (Table 3, Table S4). In the mixed model, relationship of the deceased person to the bereaved showed strong differences in IOV, with close relationships having significantly (*P* = 0.002) higher IOV than more distant relationships.

##### Age of the deceased

The age of the deceased person appeared to have a distinct effect on levels of grief and perceived support needs; in particular, there were distinct trends of reductions in most of these outcomes for ages above 40 to 50 years old (Figure 1). This negative trend in IOV with age was also apparent in the mixed model (not shown here).

**Figure 1:**
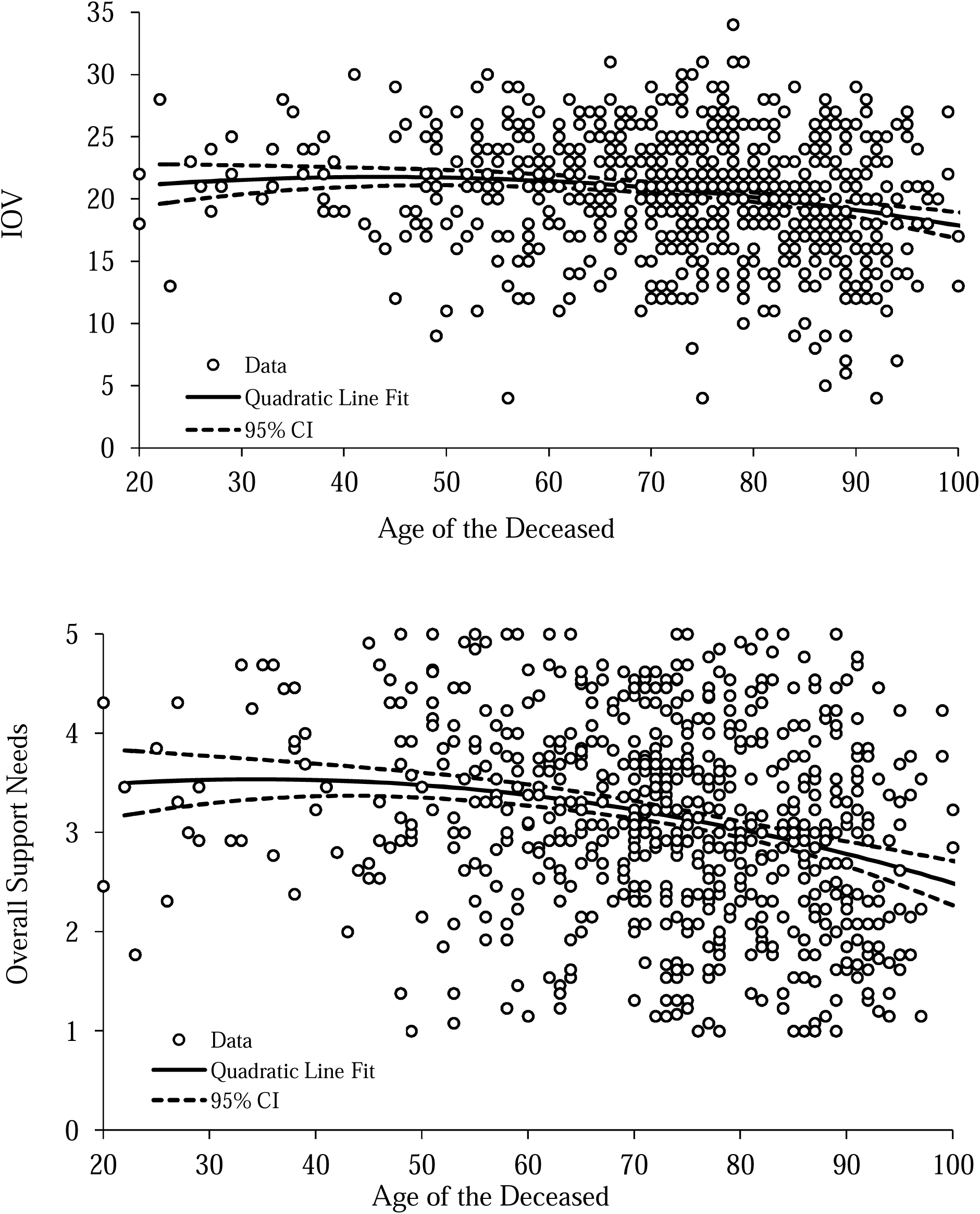
Scatter plots showing levels of grief (IOV) and overall support score as a function of the age of the person who died. Quadratic line fits and associated 95% confidence intervals on the estimate have been added to these figures to show the general trend more clearly, namely, of a distinct reduction in these outcomes with age above 40 to 50 years.

##### Social isolation and loneliness

Bereaved participants who experienced social isolation and loneliness experienced significantly higher (*P* < 0.001) levels of grief and needed more support (especially emotional support, where a large effect size was observed) than those who did not (Table 4). Overall, 58% of those who experienced social isolation/loneliness reported high or severe levels of vulnerability as measured by IOV, compared with 38.4% of those who did not. The absolute measure of effect size via a difference in these percentages is 19.6% (Cohen’s *h* = 0.46).

**Table 4:** Results for subscales and scale scores for the AAG questionnaire and support needed for social isolation and loneliness (No / Yes)

|  |  | AAG |  |  |  | Support Needed |  |  |
| --- | --- | --- | --- | --- | --- | --- | --- | --- |
|  |  | Overwhelmed | Controlled | Reversed Resilience | IOV | Practical | Emotional | Overall |
| No | *n* | 233 | 230 | 231 | 229 | 227 | 228 | 228 |
|  | Mean | 7.62 | 6.87 | 4.49 | 18.99 | 2.07 | 2.74 | 2.59 |
|  | Median | 8 | 7 | 4 | 19 | 2 | 2.65 | 2.46 |
|  | SD | 2.89 | 2.6 | 2.64 | 5.07 | 0.99 | 1.10 | 0.99 |
| Yes | *n* | 472 | 470 | 470 | 469 | 463 | 468 | 468 |
|  | Mean | 8.98 | 6.49 | 5.67 | 21.11 | 2.58 | 3.62 | 3.39 |
|  | Median | 9 | 7 | 5 | 21 | 2.67 | 3.70 | 3.46 |
|  | SD | 2.634 | 2.757 | 2.821 | 4.463 | 1.11 | 0.95 | 0.89 |
| Cohen's *d* |  | 0.49 | 0.14 | 0.43 | 0.44 | 0.49 | 0.87 | 0.85 |
| *t*-test: *P* = |  | <0.001 | 0.08 | <0.001 | <0.001 | <0.001 | <0.001 | <0.001 |

##### Support from health professionals immediately after the death

Increased levels of perceived support from health professionals led to significantly (*P* < 0.001) lower levels of grief measured by IOV, the reversed resilience subscale of the AAG, and emotional and overall support need (small to medium effect, *P* < 0.001) (Table S5). This is also seen in the mixed model, where a distinct and highly significant (*P* < 0.001) decrease in IOV is seen with increasing levels of healthcare professional support immediately after the death. Other aspects of end-of-life experiences had less of an effect on outcomes and these were not significant.

##### Place of death

Place of death had an effect on all outcomes, with values for the subscale and overall scores for the AAG and support needed lower when the person died “in a care home” compared with the other groups (often *P* < 0.05), particularly for emotional and overall support, IOV and the overwhelmed AAG subscale (Table S6). A trend of reduced IOV for deaths in care homes (*P* = 0.056) compared with other places where the death occurred was also seen in the mixed model.

##### Qualifications

Highest qualification was significantly associated (*P* = 0.019) with level of grief, with more highly educated participants (with post A-level qualifications) having slightly lower IOV compared with participants with lower levels of qualification (Table S7), and this was maintained in the mixed model (*P* = 0.019).

##### Funeral restrictions

Restricted funeral arrangements also had a strong negative effect on vulnerability measured by IOV (Table S8). Experiencing other pandemic-related problems such as being unable to visit, spend time with or say goodbye to a friend or relative prior to their death had a generally small effect (and often *P* > 0.05) on levels of grief and perceived support needs (Table S8).

##### Other factors

In univariate analyses, cause of death (COVID versus non-COVID) had a small but often significant (*P* < 0.01) effect on levels of grief and perceived support needs, which were slightly higher for COVID deaths compared to non-COVID (Table S9). However, differences in IOV by cause of death were not significant (*P* = 0.66) in the mixed model. Those who did not expect their loved one to die demonstrated higher levels of grief and also support needs (again often: *P* < 0.001) (Table S10), although this was not significant in the mixed model (*P* = 0.089).

For age of the bereaved person, there was a distinct dip in the scatterplots, with levels of grief and support needs falling up to age approximately 50 and then rising (Supplementary Figure S1). Deprivation, gender, ethnicity and length of time since the death had small effects on outcomes and any observed differences were not statistically significant (*P* > 0.05) (data not shown).

## Discussion

In this national survey of people bereaved during the first nine months of the pandemic in the UK, we found that relationship to the deceased was the factor most strongly associated with higher levels of grief and support needs. Bereaved people who had lost a partner, child or sibling showed higher levels of grief and support needs compared with bereavements of more distant relatives/friends. Age of the deceased had a strong effect, with younger age a risk factor for vulnerability in grief and higher support needs. Social isolation and loneliness had a medium-large effect, particularly on emotional support needs. Other risk factors were less perceived support from health professionals after the death, place of death being in hospital, hospice or at home rather than in a care home, lower level of qualification and experiencing funeral restrictions.

The association between relationship to the deceased and higher levels of grief and support needs coheres with pre-pandemic studies (Aoun *et al*., 2015, Lobb *et al*., 2010) and studies of pandemic bereavement (Breen *et al*., 2021a, Tang and Xiang, 2021, Tang *et al*., 2021). A US survey of people bereaved by COVID-19 (*n* = 307) found a close relationship with the deceased (partner or immediate family member) was associated with higher functional impairment than a more distant relationship (extended family, friend/acquaintance) (Breen *et al*., 2021a). We found the deceased being of a younger age was associated with worse grief outcomes and experiences, which echoes other studies (Aoun *et al*., 2015, Ringdal *et al*., 2001).

The protective effects of social support, and the poor outcomes associated with loneliness, are increasingly recognised (Prohaska *et al*., 2020, Wang *et al*., 2018). Bereavement increases the risk of loneliness (Fried *et al*., 2015), while social support enables grieving people to cope (Harrop *et al*., 2020b) and protects against poor psychological outcomes (Juth *et al*., 2015, Mason *et al*., 2020), especially after traumatic deaths (Scott *et al*., 2020). Our findings support this body of research and highlight the challenges of bereavement during a time of huge disruptions to social networks. In our sample, 66.7% reported experiencing social isolation and loneliness, associated with higher vulnerability in grief and higher support needs overall (Selman *et al*., 2021b). Further, the odds of social isolation and loneliness in early bereavement were highest for bereaved partners compared with other relationships to the deceased, and in COVID-19 deaths compared with other causes of death (Selman *et al*., 2021b). Qualitative study findings demonstrate how lockdown restrictions, shielding, and mandated and self-imposed periods of isolation, prevented people from accessing the mutual comfort and support that they needed to come to terms with their loss and begin to process their grief (Torrens-Burton *et al*., 2021). This highlights the need to support people at risk of social isolation and consider the many underlying factors and sequelae.

Our finding that perceived support from healthcare professionals after death affects levels of grief and support needs demonstrates the importance of compassionate care, timely communication and support around the time of death in subsequent experiences of bereavement. This supports findings of pre-pandemic studies (Kentish-Barnes *et al*., 2015, Yamaguchi *et al*., 2017), and suggests that poor end-of-life care experiences during the pandemic (Mayland *et al*., 2021, Neimeyer and Lee, 2021) will lead to higher levels of support needs and demand on bereavement services. Similarly, our finding that lower levels of education are associated with grief vulnerability and higher support needs reflects the findings of previous research (Mason *et al*., 2020, Milic *et al*., 2017), underlining the importance of considering structural disadvantage and inequity in the assessment and provision of bereavement support. We found that outcomes were slightly better when a death had occurred in a care home compared with other settings, which may be due to anticipatory grief work, e.g. in the context of dementia diagnoses.

Pre-pandemic research regarding the impact of funerals on bereavement outcomes is inconclusive (Burrell and Selman, 2020), however even though only 7% of this sample reported not experiencing funeral restrictions, our analysis found funeral restrictions were nevertheless associated with poorer outcomes. The impact of funeral restrictions and social isolation in the context of pandemic lockdowns, social distancing and quarantining was also reflected in our qualitative data (Torrens-Burton *et al*., 2021). Participants felt that being unable to complete usual rituals, including hosting conventional services and wakes, prevented them from providing the ‘send-off’ their family member deserved, find closure and begin to grieve.

Contrary to US research (Neimeyer and Lee, 2021), we found pandemic-related problematic experiences such as being unable to visit, spend time with or say goodbye to a friend or relative prior to their death were not significantly associated with levels of grief as assessed by the AAG, or support needs. In contrast, participants gave detailed qualitative descriptions of the negative impacts that clinical and social restrictions have had on their grief (Torrens-Burton *et al*., 2021). For example, being unable to visit or say goodbye left people with feelings of intense sadness and guilt that they could not be there to comfort and support their dying relative, while some were angry as they felt the death was preventable. These findings help explain why 60% of the sample reported high/fairly high needs for help dealing with how their loved one died. It is possible the impact of some of these experiences is not fully assessed by the AAG, which aims to provide a broad grief profile of both core bereavement reactions and the coping response to them (Sim *et al*., 2014). In contrast to measures of PGD (Boelen and Smid, 2017), the AAG does not identify specific symptoms, such as longing for the dead person, anger, guilt, blame, trust in others and impact on identity and functioning, some of which may be particularly relevant in the pandemic context (Selman *et al*., 2021a). In this longitudinal study, PGD will be assessed at subsequent timepoints, reflecting the requirement that PGD is assessed at least 6 months after a death.

Five months after the death, we found high levels of vulnerability in grief overall, with 23.4% exhibiting high levels of vulnerability via the AAG (21 ≤ IOV ≤ 23), and 28.2% severe levels of vulnerability (IOV ≥ 24). Public health models of bereavement (Aoun *et al*., 2012) suggest that in non-pandemic times, 10% of bereaved people are at high risk of complex grief issues and may need referral to mental health professionals, and a further 30% are at moderate risk and may need some additional support e.g. via peer support groups. These estimates have been confirmed in an Australian study which found 6.4% at high risk and 35.2% at moderate risk (Aoun *et al*., 2015). Since acute grief is one of the strongest predictors of future disturbed grief (Boelen and Lenferink, 2020, Milic *et al*., 2017), our findings support the hypothesis that prevalence of grief disorders will rise because of the pandemic (Eisma and Boelen, 2021).

Research from the USA suggests that bereavement due to COVID-19 might be associated with elevated acute grief and post-traumatic stress, depression, and anxiety symptoms (Breen *et al*., 2021a). In China, a survey of COVID-19 bereaved adults, including a subset bereaved over six months ago, found elevated posttraumatic stress, anxiety, and depression symptoms, with between 38% and 29% meeting criteria for PGD (Tang and Xiang, 2021, Tang *et al*., 2021). In previous analyses we found that the COVID-19 bereaved had worse experiences of end-of-life and early bereavement, including higher rates of social isolation and loneliness (Selman *et al*., 2021b). In the current analysis COVID-19 resulted in a mild increase in outcomes (including severity of grief) in univariate calculations, although this was not significant in the mixed model for IOV. Analyses of our longitudinal study data will help explicate the mechanisms and impact at play here.

The higher levels of grief observed in this sample compared with pre-pandemic studies may be due to stressors universally experienced during the pandemic rather than specific to COVID-19 bereavements, including disruptions to the meaning-making process following a death (Breen *et al*., 2021b). (Eisma and Tamminga, 2020) found that people in the Netherlands who recently experienced a non-COVID-19 death during the pandemic reported higher levels of acute grief than those recently bereaved before the pandemic. (Breen *et al*., 2021b) found that among people bereaved during the COVID-19 pandemic in the USA (N=409), there were no statistically significant differences in grief outcomes or functional impairment according to cause of death. Further research is required to establish which experiences and symptomatology are specific to the COVID-19 bereaved and which apply to the broader population of people bereaved during the pandemic.

### Strengths and weaknesses

The study sample was large, with good spread across geographical areas, education and deprivation, but relied on voluntary response sampling. The sample was biased towards female and white respondents, despite targeted advertising to men and people from ethnic minority communities. By recruiting mostly online, we were less likely to reach the very old or other digitally marginalised groups. Through subsequent qualitative interviews, we explore in depth the experiences of people from communities and groups less well represented in the survey. Convenience sampling might have resulted in more people with negative experiences completing the survey. Despite these limitations, group sizes were sufficient to enable comparisons (although not to the level of specific ethnic groups) and, while not providing population-level prevalence data, the sample does enable identification of risk factors to inform future practice and policy.

### Implications for research

This study includes follow-up surveys at c.7 and 13 months post-death, longitudinal qualitative interviews and research exploring the impact of the pandemic on voluntary sector bereavement services and their response. Further research exploring the needs of bereaved people from minority ethnic backgrounds, same-sex couple, men, children and young people and longer-term bereavement experiences is also required, particularly population-based studies to establish the prevalence of PGD and other poor outcomes among the pandemic-bereaved over time.

### Conclusions and implications for policy and practice

Study findings inform bereavement support and policy during and beyond this and future pandemics. Results highlight the particular complexities and challenges of pandemic bereavement as well as identifying who is at particular risk of poor grief outcomes and may have higher levels of support need. We make the following evidence-based recommendations:

1. Given high levels of grief vulnerability and needs for bereavement support, especially psycho-emotional support, among people bereaved during the COVID-19 pandemic, statutory, voluntary and community bereavement support services require increased investment, underpinned by national and local policies to meet high levels of need.
2. Close relatives are at particular risk of poor outcomes, especially when socially isolated, and should be targeted for additional support and follow-up after a death.
3. The importance of funerals and other group mourning social practices must be recognised, with restrictions considered carefully. Funeral providers and celebrants play an important role in providing meaningful services in the contexts of restrictions (Burrell and Selman, 2020).
4. The quality of care and support provided to bereaved people immediately after a death influences bereavement outcomes and support needs and must be prioritised and adequately resourced in the pandemic context, just as in non-pandemic times.
5. Clinicians and bereavement services need to consider the needs of and prioritise people less able to advocate for themselves, including those subject to structural disadvantage and inequity.
6. Following public health strategies, compassionate community-based initiatives in bereavement are needed to strengthen, support and learn from communities’ own approaches to informal bereavement support.

## Supporting information

CHERRIES reporting checklist

## Data Availability

All data produced in the present study are available upon reasonable request to the authors

## Acknowledgements

Our thanks to everyone who completed the survey for sharing their experiences, and to all the individuals and organisations that helped disseminate the survey. We would also like to thank the project assistants, collaborators and advisory group members who are not co-authors on this publication: Dr. Emma Carduff, Dr Daniella Holland-Hart, Prof. Bridget Johnston, Dr. Donna Wakefield, Alison Penny, Dr Kirsten Smith, Dr. Audrey Roulston and Dr. Anne Finucane.

## Ethical standards

The authors assert that all procedures contributing to this work comply with the ethical standards of the relevant national and institutional committees on human experimentation and with the Helsinki Declaration of 1975, as revised in 2008.

## Data availability

Full study data sets will be made available following study closure in February 2022. Data sharing requests will be considered prior to this and should be directed to Dr. Emily Harrop.

## References

Aoun, S. M., Breen, L. J., Howting, D. A., Rumbold, B., McNamara, B. & Hegney, D. (2015). Who needs bereavement support? A population based survey of bereavement risk and support need. PLoS ONE 10, e0121101.

Aoun, S. M., Breen, L. J., O’ Connor, M., Rumbold, B. & Nordstrom, C. (2012). A public health approach to bereavement support services in palliative care. Aust N Z J Public Health 36, 14–6.

Boelen, P. A. & Lenferink, L. I. M. (2020). Symptoms of prolonged grief, posttraumatic stress, and depression in recently bereaved people: symptom profiles, predictive value, and cognitive behavioural correlates. Soc Psychiatry Psychiatr Epidemiol 55, 765–777.

Boelen, P. A. & Smid, G. E. (2017). The Traumatic Grief Inventory Self-Report Version (TGI-SR): Introduction and Preliminary Psychometric Evaluation. Journal of Loss and Trauma 22, 196–212.

Breen, L. J., Lee, S. A. & Neimeyer, R. A. (2021a). Psychological Risk Factors of Functional Impairment After COVID-19 Deaths. J Pain Symptom Manage 61, e1–e4.

Breen, L. J., Mancini, V. O., Lee, S. A., Pappalardo, E. A. & Neimeyer, R. A. (2021b). Risk factors for dysfunctional grief and functional impairment for all causes of death during the COVID-19 pandemic: The mediating role of meaning. Death Stud, 1–10.

Burrell, A. & Selman, L. E. (2020). How do Funeral Practices impact Bereaved Relatives’ Mental Health, Grief and Bereavement? A Mixed Methods Review with Implications for COVID-19. OMEGA-Journal of Death and Dying, 0030222820941296.

Claessen, S. J. J., Francke, A. L., Sixma, H. J., de Veer, A. J. E. & Deliens, L. (2013). Measuring Relatives’ Perspectives on the Quality of Palliative Care: The Consumer Quality Index Palliative Care. Journal of Pain and Symptom Management 45, 875–884.

Eisma, M. C. & Boelen, P. A. (2021). Commentary on: A Call to Action: Facing the Shadow Pandemic of Complicated Forms of Grief. OMEGA - Journal of Death and Dying 0, 00302228211016227.

Eisma, M. C. & Tamminga, A. (2020). Grief Before and During the COVID-19 Pandemic: Multiple Group Comparisons. J Pain Symptom Manage 60, e1–e4.

Eisma, M. C., Tamminga, A., Smid, G. E. & Boelen, P. A. (2021). Acute grief after deaths due to COVID-19, natural causes and unnatural causes: An empirical comparison. Journal of Affective Disorders 278, 54–56.

Eysenbach, G. (2004). Improving the quality of Web surveys: the Checklist for Reporting Results of Internet E-Surveys (CHERRIES). Journal of medical Internet research 6, e34–e34.

Fried, E. I., Bockting, C., Arjadi, R., Borsboom, D., Amshoff, M., Cramer, A. O., Epskamp, S., Tuerlinckx, F., Carr, D. & Stroebe, M. (2015). From loss to loneliness: The relationship between bereavement and depressive symptoms. J Abnorm Psychol 124, 256–65.

Hanna, J. R., Rapa, E., Dalton, L. J., Hughes, R., McGlinchey, T., Bennett, K. M., Donnellan, W. J., Mason, S. R. & Mayland, C. R. (2021). A qualitative study of bereaved relatives’ end of life experiences during the COVID-19 pandemic. Palliative Medicine 35, 843–851.

Harrop, E., Goss, S., Farnell, D., Longo, M., Byrne, A., Barawi, K., Torrens-Burton, A., Nelson, A., Seddon, K., Machin, L., Sutton, E., Roulston, A., Finucane, A., Penny, A., Smith, K. V., Sivell, S. & Selman, L. E. (2021). Support needs and barriers to accessing support: Baseline results of a mixed-methods national survey of people bereaved during the COVID-19 pandemic. Palliative Medicine In Press.

Harrop, E., Morgan, F., Byrne, A. & Nelson, A. (2016). “It still haunts me whether we did the right thing”: a qualitative analysis of free text survey data on the bereavement experiences and support needs of family caregivers. BMC Palliative Care 15, 92.

Harrop, E., Morgan, F., Longo, M., Semedo, L., Fitzgibbon, J., Pickett, S., Scott, H., Seddon, K., Sivell, S., Nelson, A. & Byrne, A. (2020a). The impacts and effectiveness of support for people bereaved through advanced illness: A systematic review and thematic synthesis. Palliative Medicine 34, 871–888.

Harrop, E., Scott, H., Sivell, S., Seddon, K., Fitzgibbon, J., Morgan, F., Pickett, S., Byrne, A., Nelson, A & Longo, M. (2020b). Coping and wellbeing in bereavement: two core outcomes for evaluating bereavement support in palliative care. BMC Palliative Care 19, 29.

Harrop E. F.D., Longo M., Goss S., Sutton E., Seddon K., Nelson A., Byrne A., Selman L.E. (2020). Supporting people bereaved during COVID-19: Study Report 1.

Haugen, D. F., Hufthammer, K. O., Gerlach, C., Sigurdardottir, K., Hansen, M. I. T., Ting, G., Tripodoro, V. A., Goldraij, G., Yanneo, E. G., Leppert, W., Wolszczak, K., Zambon, L., Passarini, J. N., Saad, I. A. B., Weber, M., Ellershaw, J. & Mayland, C. R. (2021). Good Quality Care for Cancer Patients Dying in Hospitals, but Information Needs Unmet: Bereaved Relatives’ Survey within Seven Countries. Oncologist 26, e1273–e1284.

JISC (2021). JISC Online Surveys.

Juth, V., Smyth, J. M., Carey, M. P. & Lepore, S. J. (2015). Social Constraints are Associated with Negative Psychological and Physical Adjustment in Bereavement. Applied Psychology: Health and Well-Being 7, 129–148.

Kentish-Barnes, N., Chaize, M., Seegers, V., Legriel, S., Cariou, A., Jaber, S., Lefrant, J.-Y., Floccard, B, Renault, A., Vinatier, I., Mathonnet, A., Reuter, D., Guisset, O., Cohen-Solal, Z., Cracco, C., Seguin, A., Durand-Gasselin, J., Éon, B., Thirion, M., Rigaud, J.-P., Philippon-Jouve, B., Argaud, L., Chouquer, R., Adda, M., Dedrie, C., Georges, H., Lebas, E., Rolin, N., Bollaert, P.-E., Lecuyer, L., Viquesnel, G., Léone, M., Chalumeau-Lemoine, L., Garrouste, M., Schlemmer, B., Chevret, S., Falissard, B. & Azoulay, É. (2015). Complicated grief after death of a relative in the intensive care unit. European Respiratory Journal 45, 1341.

Kristensen, P., Weisæth, L. & Heir, T. (2012). Bereavement and Mental Health after Sudden and Violent Losses: A Review. Psychiatry: Interpersonal and Biological Processes 75, 76–97.

Lobb, E. A., Kristjanson, L. J., Aoun, S. M., Monterosso, L., Halkett, G. K. & Davies, A. (2010). Predictors of complicated grief: a systematic review of empirical studies. Death Stud 34, 673–98.

Machin, L. (2001). Exploring a framework for understanding the range of response to loss; a study of clients receiving bereavement counselling.. Keele University, UK.

Machin, L. (2013). Working with Loss and Grief: A Theoretical and Practical Approach (2nd edition). Sage Publications Ltd. : London.

Mason, T. M., Tofthagen, C. S. & Buck, H. G. (2020). Complicated Grief: Risk Factors, Protective Factors, and Interventions. Journal of Social Work in End-of-Life & Palliative Care 16, 151–174.

Mayland, C. R., Hughes, R., Lane, S., McGlinchey, T., Donnellan, W., Bennett, K., Hanna, J., Rapa, E., Dalton, L. & Mason, S. R. (2021). Are public health measures and individualised care compatible in the face of a pandemic? A national observational study of bereaved relatives’ experiences during the COVID-19 pandemic. Palliative Medicine 0, 02692163211019885.

Milic, J., Muka, T., Ikram, M. A., Franco, O. H. & Tiemeier, H. (2017). Determinants and Predictors of Grief Severity and Persistence: The Rotterdam Study. J Aging Health 29, 1288–1307.

Miyashita, M., Morita, T., Sato, K., Hirai, K., Shima, Y. & Uchitomi, Y. (2008). Factors contributing to evaluation of a good death from the bereaved family member’s perspective. Psychooncology 17, 612–20.

Neimeyer, R. A. & Lee, S. A. (2021). Circumstances of the death and associated risk factors for severity and impairment of COVID-19 grief. Death Stud, 1–9.

Prohaska, T., Burholt, V., Burns, A., Golden, J., Hawkley, L., Lawlor, B., Leavey, G., Lubben, J., O’ Sullivan, R., Perissinotto, C., van Tilburg, T., Tully, M., Victor, C. & Fried, L. (2020). Consensus statement: loneliness in older adults, the 21st century social determinant of health? BMJ Open 10, e034967.

Ringdal, G. I., Jordhøy, M. S., Ringdal, K. & Kaasa, S. (2001). Factors Affecting Grief Reactions in Close Family Members to Individuals Who Have Died of Cancer. Journal of Pain and Symptom Management 22, 1016–1026.

Scott, H. R., Pitman, A., Kozhuharova, P. & Lloyd-Evans, B. (2020). A systematic review of studies describing the influence of informal social support on psychological wellbeing in people bereaved by sudden or violent causes of death. BMC Psychiatry 20, 265.

Selman, L. E., Chamberlain, C., Sowden, R., Chao, D., Selman, D., Taubert, M. & Braude, P. (2021a). Sadness, despair and anger when a patient dies alone from COVID-19: A thematic content analysis of Twitter data from bereaved family members and friends. Palliative Medicine 0, 02692163211017026.

Selman, L. E., Chao, D., Sowden, R., Marshall, S., Chamberlain, C. & Koffman, J. (2020). Bereavement support on the frontline of COVID-19: Recommendations for hospital clinicians. Journal of Pain and Symptom Management 60, e81–e86.

Selman, L. E., Farnell, D., Longo, M., Goss, S., Seddon, K., Torrens-Burton, A., Mayland, C. R., Wakefield, D., Johnston, B., Byrne, A. & Harrop, E. (2021b). Risk factors associated with poorer experiences of end-of-life care and challenges in early bereavement: Results of a national online survey of people bereaved during the COVID-19 pandemic. Palliat Med In Press.

Sim, J., Machin, L. & Bartlam, B. (2014). Identifying vulnerability in grief: psychometric properties of the Adult Attitude to Grief Scale. Qual Life Res 23, 1211–20.

Smith, K. V., Wild, J. & Ehlers, A. (2020). The Masking of Mourning: Social Disconnection After Bereavement and Its Role in Psychological Distress. Clinical Psychological Science 8, 464–476.

Sue Ryder (2019). A Better Grief Sue Ryder: London, UK.

Tang, S. & Xiang, Z. (2021). Who suffered most after deaths due to COVID-19? Prevalence and correlates of prolonged grief disorder in COVID-19 related bereaved adults. Globalization and Health 17, 19.

Tang, S., Yu, Y., Chen, Q., Fan, M. & Eisma, M. C. (2021). Correlates of Mental Health After COVID-19 Bereavement in Mainland China. Journal of Pain and Symptom Management 61, e1–e4.

Torrens-Burton, A., Goss, S., Sutton, E., Barawi, K., Longo, M., Seddon, K., Carduff, E., Farnell, D., Nelson, A., Byrne, A., Selman, L. E. & Harrop, E. (2021). ‘It was brutal. It still is’ : A qualitative analysis of the challenges of bereavement during the COVID-19 pandemic reported in two national surveys [preprint]. medRxiv.

Verdery, A. M., Smith-Greenaway, E., Margolis, R. & Daw, J. (2020). Tracking the reach of COVID-19 kin loss with a bereavement multiplier applied to the United States. Proceedings of the National Academy of Sciences 117, 17695–17701.

Wang, J., Mann, F., Lloyd-Evans, B., Ma, R. & Johnson, S. (2018). Associations between loneliness and perceived social support and outcomes of mental health problems: a systematic review. BMC Psychiatry 18, 156.

Yamaguchi, T., Maeda, I., Hatano, Y., Mori, M., Shima, Y., Tsuneto, S., Kizawa, Y., Morita, T., Yamaguchi, T., Aoyama, M. & Miyashita, M. (2017). Effects of End-of-Life Discussions on the Mental Health of Bereaved Family Members and Quality of Patient Death and Care. Journal of Pain and Symptom Management 54, 17–26.e1.

Yardley, S. & Rolph, M. (2020). Death and dying during the pandemic. BMJ 369, m1472.

