## Supplementary file 2 for "Factors associated with higher levels of grief and support needs among people bereaved during the pandemic: Results from a national online survey"

Supplementary file 2: Survey advertisement

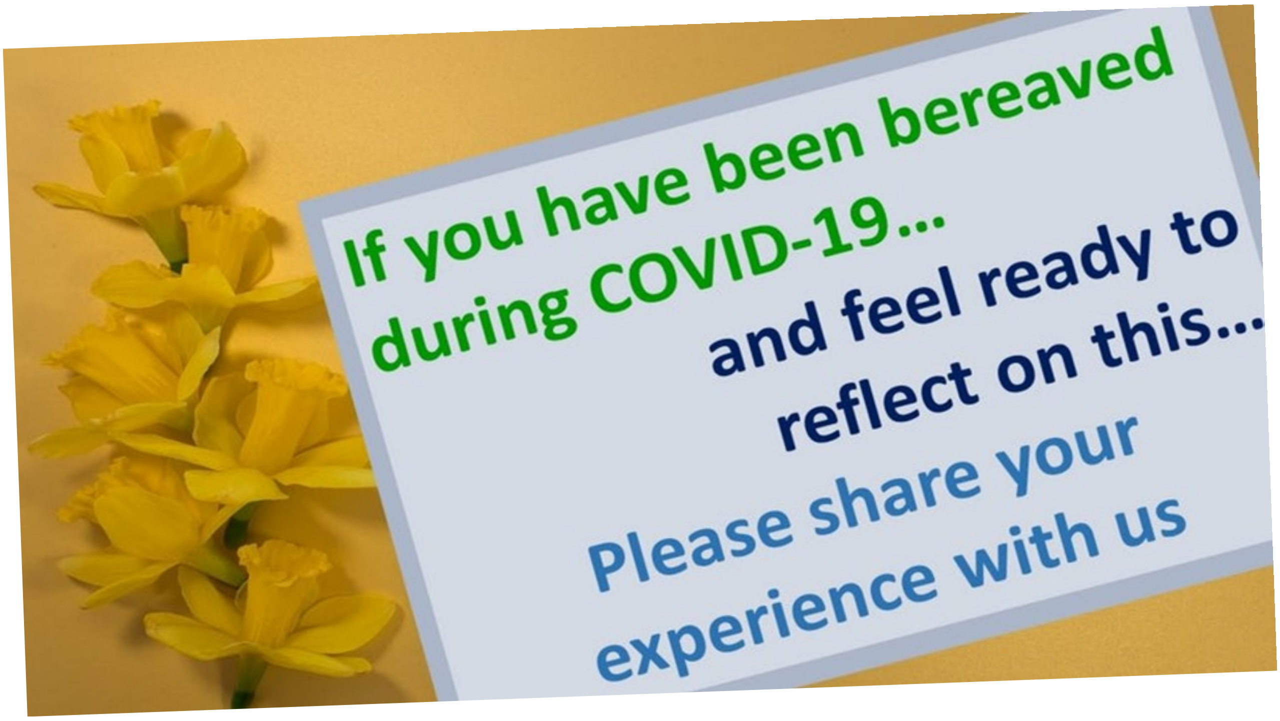


Cardiff University and the University of Bristol are conducting a **survey** looking at the **grief experiences** and **support needs** of people **bereaved** during the **pandemic.**

By conducting this survey we hope to identify ways of improving the care provided at the end of life and during bereavement.

If you have lost a loved one to COVID-19 or another cause of death during the pandemic, and would like to share your experience in our survey, please **click here** or visit www.covidbereavement.com

Or contact Emily Harrop: ****, **tel: 02920 687184** for further information or a paper copy of the survey.

### Thank you!

Cardiff University / Prifysgol Caerdydd · ESRC Economic & Social Research Council · University of Bristol · Marie Curie · Canolfan Ymchwil Gofal Lliniarol Marie Curie Palliative Care Research Centre Cardiff Caerdydd
