## Supplementary file 3 Tables for "Factors associated with higher levels of grief and support needs among people bereaved during the pandemic: Results from a national online survey"

**Supplementary Tables and Figures**

**S1: Frequency table for items in the AAG questionnaire (n = 711)**

| **AAG item (and score)** | | **4 = Strongly agree**  **n (%)** | **3 = Agree**  **n (%)** | **2= Neither agree or disagree**  **n (%)** | **1= Disagree n (%)** | **0 = Strongly disagree**  **n (%)** | **Missing**  **n (%)** | **Mean**  **(95% CI)** | **Median** |
| --- | --- | --- | --- | --- | --- | --- | --- | --- | --- |
| Overwhelmed items | 2. For me, it is difficult to switch off thoughts about the person I have lost. | 309 (43.5) | 217 (30.5) | 95 (13.4) | 58 (8.2) | 27 (3.8) | 5 (0.7) | 3.02 (2.93 to 3.1) | 3 |
|  | 5. I feel that I will always carry the pain of grief with me. | 330 (46.4) | 230 (32.3) | 79 (11.1) | 47 (6.6) | 18 (2.5) | 7 (1) | 3.15 (3.07 to 3.23) | 3 |
|  | 7. Life has less meaning for me after this loss. | 174 (24.5) | 172 (24.2) | 157 (22.1) | 129 (18.1) | 69 (9.7) | 10 (1.4) | 2.36 (2.26 to 2.45) | 2 |
| Controlled items | 4. I believe that I must be brave in the face of loss. | 102 (14.3) | 266 (37.4) | 151 (21.2) | 126 (17.7) | 57 (8) | 9 (1.3) | 2.33 (2.24 to 2.42) | 3 |
|  | 6. For me, it is important to keep my grief under control. | 93 (13.1) | 284 (39.9) | 164 (23.1) | 107 (15) | 52 (7.3) | 11 (1.5) | 2.37 (2.28 to 2.45) | 3 |
|  | 8. I think it’s best just to get on with life in spite of this loss. | 44 (6.2) | 200 (28.1) | 203 (28.6) | 153 (21.5) | 97 (13.6) | 14 (2) | 1.92 (1.82 to 1.99) | 2 |
| **AAG item (and score)** | | **0 = Strongly agree**  **n (%)** | **1 = Agree**  **n (%)** | **2 = Neither agree or disagree**  **n (%)** | **3 = Disagree n (%)** | **4 Strongly disagree**  **n (%)** | **Missing**  **n (%)** | **Mean**  **(95% CI)** | **Median** |
|  | 1. I feel able to face the pain which comes with loss. | 55 (7.7) | 228 (32.1) | 126 (17.7) | 173 (24.3) | 116 (16.3) | 13 (1.8) | 2.1 (2.00 to 2.19) | 2 |
| Reversed Resilience Items | 3. I feel very aware of my inner strength when faced with grief. | 88 (12.4) | 225 (31.6) | 202 (28.4) | 141 (19.8) | 45 (6.3) | 10 (1.4) | 1.76 (1.67 to 1.84) | 2 |
|  | 9. It may not always feel like it but I do believe that I will come through this experience of grief. | 111 (15.6) | 333 (46.8) | 145 (20.4) | 73 (10.3) | 39 (5.5) | 10 (1.4) | 1.42 (1.35 to 1.51) | 1 |

**Table S2: Descriptive statistics for the AAG questionnaire**

| Subscales/  Total score | *n* | % Missing | Mean (95% CI) [Median] | SD |
| --- | --- | --- | --- | --- |
| Overwhelmed | 705 | 0.8% | 8.53 (8.31 to 8.72) [9.00] | 2.79 |
| Controlled | 700 | 1.5% | 6.61 (6.41 to 6.82) [7.00] | 2.71 |
| Reversed Resilience | 701 | 1.4% | 5.28 (5.07 to 5.49) [5.00] | 2.82 |
| IOV | 698 | 1.8% | 20.41 (20.06 to 20.77) [21.00] | 4.77 |
| IOV Categories | *n (%)* |  | | |
| Low (IOV = 0-20) | 338 (48.4) |  |  |  |
| High (IOV = 21-23) | 163 (23.4) |  |  |  |
| Severe (IOV = 24-36) | 197 (28.2) |  |  |  |

**Table S3: Support needs ranked by mean level of need**

|  | High or fairly high level of support needed | Moderate level of support needed | Little or no support needed | Mean  (95% CI) | Median |
| --- | --- | --- | --- | --- | --- |
| Dealing with my feelings about the way my loved one died | 59.8% | 21.5% | 18.7% | 3.71  (3.62 to 3.80) | 4 |
| Dealing with my feelings about being without my loved one | 49.9% | 29.3% | 20.8% | 3.48  (3.39 to 3.57) | 3 |
| Expressing my feelings and feeling understood by others | 53% | 23.9% | 23% | 3.48  (3.38 to 3.57) | 4 |
| Feeling comforted and reassured | 51.8% | 26.7% | 21.6% | 3.46  (3.37 to 3.55) | 4 |
| Feelings of anxiety and depression | 52.8% | 21.1% | 26.1% | 3.45  (3.35 to 3.55) | 4 |
| Loneliness and social isolation | 52.0% | 19.1% | 29% | 3.36  (3.26 to 3.46) | 4 |
| Finding balance between grieving and other areas of life | 45.0% | 27.9% | 27% | 3.29  (3.20 to 3.39) | 3 |
| Regaining sense of purpose and meaning in life | 46.7% | 21.6% | 31.7% | 3.26  (3.15 to 3.36) | 3 |
| Managing and maintaining my relationships with friends and family | 36.2% | 26.4% | 37.4% | 2.98  (2.88 to 3.08) | 3 |
| Participating in work, leisure or other regular activities (e.g. shopping, housework) | 33.8% | 23.9% | 42.1% | 2.87  (2.76 to 2.97) | 3 |
| Getting relevant information and advice e.g. legal, financial, available support | 24.3% | 22.3% | 53.3% | 2.51  (2.41 to 2.61) | 2 |
| Practical tasks e.g. managing the funeral, registering the death, other paperwork etc. | 23.5% | 21.7% | 54.7% | 2.48  (2.38 to 2.58) | 2 |
| Looking after myself/family e.g. getting food, medication, childcare | 15.2% | 22.8% | 62% | 2.25  (2.16 to 2.34) | 2 |

Note to interpret means and medians: no support = 1; little support = 2; moderate support = 3; fairly high support = 4; and high support = 5

**Table S4: Results for the three IOV groups (i.e., low, high, & extreme) by relationship of bereaved person to the deceased. (Overall association between IOV and relationship: Chi-squared test: *P* < 0.001; “row” percentages shown are shown below, i.e., with respect to *n* quoted in each row for each type of relationship to the deceased.)**

|  | *n* | Low | High | Severe |
| --- | --- | --- | --- | --- |
| Partners | 148 | 34.5% | 28.4% | 37.2% |
| Parents | 390 | 50.3% | 21.0% | 28.7% |
| Grandparents | 54 | 57.4% | 27.8% | 14.8% |
| Sibling | 22 | 36.4% | 40.9% | 22.7% |
| Child | 15 | 20.0% | 40.0% | 40.0% |
| Other family member | 45 | 68.9% | 15.6% | 15.6% |
| Colleague or friend | 24 | 75.0% | 8.3% | 16.7% |

**Table S5: Results for subscales and overall scale scores for the AAG questionnaire and support needed as a function of “Did you feel well supported by the healthcare professionals immediately after the death of your loved one?”**

|  | | AAG | | | | Support Needed | | |
| --- | --- | --- | --- | --- | --- | --- | --- | --- |
|  | | Overwhelmed | Controlled | Reversed Resilience | IOV | Practical | Emotional | Overall |
| Very well supported | *n* | 95 | 95 | 95 | 95 | 93 | 93 | 93 |
|  | Mean | 8.21 | 6.06 | 4.42 | 18.69 | 2.40 | 3.06 | 2.90 |
|  | SD | 2.58 | 2.74 | 2.66 | 4.50 | 1.00 | 1.04 | 0.96 |
|  | Median | 8 | 6 | 4 | 20 | 2.33 | 3.1 | 2.92 |
| Fairly well supported | *N* | 105 | 103 | 103 | 103 | 105 | 104 | 104 |
|  | Mean | 8.72 | 6.93 | 5.14 | 20.73 | 2.43 | 3.31 | 3.10 |
|  | SD | 2.88 | 2.72 | 2.56 | 4.78 | 1.06 | 1.05 | 0.96 |
|  | Median | 9 | 7 | 5 | 21 | 2.33 | 3.35 | 3.08 |
| A little bit supported | *n* | 138 | 137 | 139 | 137 | 136 | 136 | 136 |
|  | Mean | 8.77 | 6.63 | 5.1 | 20.46 | 2.54 | 3.39 | 3.20 |
|  | SD | 2.60 | 2.54 | 2.71 | 4.55 | 1.07 | 1.02 | 0.94 |
|  | Median | 9 | 7 | 5 | 21 | 2.5 | 3.55 | 3.23 |
| Not at all supported | *n* | 250 | 248 | 247 | 246 | 251 | 249 | 249 |
|  | Mean | 9.11 | 6.41 | 5.93 | 21.46 | 2.61 | 3.59 | 3.36 |
|  | SD | 2.71 | 2.77 | 2.95 | 4.62 | 1.12 | 1.05 | 0.96 |
|  | Median | 10 | 7 | 6 | 22 | 2.33 | 3.7 | 3.38 |
| Maximum Cohen’s \|*d*\| | | 0.33 | 0.32 | 0.56 | 0.60 | 0.20 | 0.50 | 0.47 |
| One-way ANOVA: *P* = | | 0.05 | 0.13 | <0.001 | <0.001 | 0.28 | <0.001 | 0.001 |

**Table S6: Results for subscales and scale scores for the AAG questionnaire and support needed as a function of place of death**

|  | | AAG | | | | Support Needed | | |
| --- | --- | --- | --- | --- | --- | --- | --- | --- |
|  | | Overwhelmed | Controlled | Reversed Resilience | IOV | Practical | Emotional | Overall |
| In hospital | *n* | 406 | 404 | 406 | 403 | 395 | 400 | 400 |
|  | Mean | 8.84 | 6.45 | 5.52 | 20.79 | 2.51 | 3.44 | 3.23 |
|  | SD | 2.76 | 2.76 | 2.92 | 4.64 | 1.14 | 1.05 | 0.97 |
|  | Median | 9 | 7 | 5 | 21 | 2.33 | 3.55 | 3.23 |
| In their home | *N* | 157 | 156 | 156 | 156 | 152 | 154 | 154 |
|  | Mean | 8.31 | 7.01 | 5.04 | 20.34 | 2.40 | 3.24 | 3.05 |
|  | SD | 2.74 | 2.50 | 2.81 | 4.85 | 1.10 | 1.15 | 1.06 |
|  | Median | 9 | 7 | 5 | 21 | 2.33 | 3.2 | 3 |
| In a hospice | *n* | 37 | 35 | 35 | 35 | 37 | 37 | 37 |
|  | Mean | 8.86 | 6.94 | 5.20 | 20.83 | 2.24 | 3.36 | 3.10 |
|  | SD | 2.69 | 2.86 | 2.56 | 5.46 | 0.95 | 0.89 | 0.81 |
|  | Median | 8 | 8 | 5 | 21 | 2 | 3.3 | 3 |
| In a care home | *n* | 90 | 90 | 89 | 89 | 91 | 90 | 90 |
|  | Mean | 7.34 | 6.68 | 4.65 | 18.70 | 2.08 | 2.96 | 2.75 |
|  | SD | 2.85 | 2.67 | 2.35 | 4.78 | 0.88 | 1.06 | 0.96 |
|  | Median | 7.5 | 7 | 4 | 19 | 2 | 3 | 2.77 |
| Other / Do not Know | *n* | 13 | 13 | 13 | 13 | 13 | 13 | 13 |
|  | Mean | 9.08 | 5.31 | 5.08 | 19.46 | 2.51 | 3.57 | 3.32 |
|  | SD | 2.47 | 3.04 | 2.75 | 3.80 | 1.16 | 1.37 | 1.22 |
|  | Median | 9 | 6 | 5 | 19 | 2.33 | 4 | 3.62 |
| Cohen’s *d* | | 0.64 | 0.61 | 0.32 | 0.45 | 0.41 | 0.55 | 0.57 |
| Maximum Cohen’s \|*d*\| | | <0.001 | 0.10 | 0.05 | 0.01 | 0.01 | 0.01 | 0.003 |
| One-way ANOVA: *P* = | | <0.001 | 0.158 | 0.11 | 0.002 | 0.036 | 0.002 | 0.001 |

**Table S7: Results for subscales and scale scores for the AAG questionnaire and support needed as a function of highest qualification.**

|  | | AAG | | | | Support Needed | | |
| --- | --- | --- | --- | --- | --- | --- | --- | --- |
|  | | Overwhelmed | Controlled | Reversed Resilience | IOV | Practical | Emotional | Overall |
| None / GCSEs | *n* | 103 | 101 | 102 | 100 | 104 | 104 | 104 |
|  | Mean | 9.48 | 5.94 | 6.42 | 21.77 | 2.63 | 3.52 | 3.32 |
|  | SD | 3.00 | 2.83 | 2.94 | 4.1 | 1.19 | 1.07 | 1.02 |
|  | Median | 10 | 6 | 6.5 | 22 | 2.67 | 3.68 | 3.40 |
| A-level/ apprenticeship / ONC | *n* | 132 | 132 | 131 | 131 | 127 | 129 | 129 |
|  | Mean | 9.29 | 6.61 | 5.55 | 21.48 | 2.53 | 3.57 | 3.33 |
|  | SD | 2.56 | 2.67 | 2.84 | 4.52 | 1.08 | 1.06 | 0.98 |
|  | Median | 10 | 7 | 6 | 22 | 2.33 | 3.8 | 3.38 |
| HND / University Degree / Postgraduate (etc) | *n* | 467 | 464 | 465 | 464 | 456 | 460 | 460 |
|  | Mean | 8.11 | 6.76 | 4.94 | 19.8 | 2.32 | 3.22 | 3.02 |
|  | SD | 2.71 | 2.68 | 2.70 | 4.86 | 1.06 | 1.07 | 0.98 |
|  | Median | 8 | 7 | 5 | 20 | 2 | 3.25 | 3 |
| Maximum Cohen’s \|*d*\| | | 0.50 | 0.30 | 0.52 | 0.44 | 0.28 | 0.33 | 0.32 |
| One-way ANOVA: *P* = | | <0.001 | 0.03 | <0.001 | <0.001 | 0.02 | 0.001 | 0.001 |

**Table S8: Percentages for the 3 groups with respect to IOV (low, high, severe) for pandemic-related problems.**

|  | | *n* = | IOV Group | | | Cohen’s *h* | *P* = |
| --- | --- | --- | --- | --- | --- | --- | --- |
|  |  |  | Low | High | Severe |  |  |
| Unable to visit them prior to their death | No | 320 | 49.7% | 24.4% | 25.9% | 0.088 | 0.459 |
|  | Yes | 378 | 47.4% | 22.5% | 30.2% |  |  |
| Limited contact with them in last days of their life | No | 293 | 47.1% | 25.3% | 27.6% | 0.068 | 0.596 |
|  | Yes | 405 | 49.4% | 22.0% | 28.6% |  |  |
| Unable to say goodbye as I would have liked | No | 253 | 49.8% | 23.3% | 26.9% | 0.050 | 0.811 |
|  | Yes | 445 | 47.6% | 23.4% | 29.0% |  |  |
| Restricted funeral arrangements^†^ | No | 45 | 66.7% | 8.9% | 24.4% | 0.477 | 0.018 |
|  | Yes | 653 | 47.2% | 24.3% | 28.5% |  |  |
| Social isolation and loneliness | No | 229 | 61.6% | 19.7% | 18.8% | 0.460 | <0.001 |
|  | Yes | 469 | 42.0% | 25.2% | 32.8% |  |  |
| Limited contact with other close relatives or friends | No | 131 | 48.9% | 23.7% | 27.5% | 0.019 | 0.994 |
|  | Yes | 567 | 48.3% | 23.3% | 28.4% |  |  |

Note: The same sizes for the responses “no” and “yes” are shown for each question. Results for Cohen’s *h* measure of effect size between two proportions indicates zero to small effect sizes only. *P*-values are from chi-squared analysis. “Row” percentages shown are shown below, i.e., with respect to *n* quoted in each row for yes / no answers for each type of pandemic related problem.

^†^Sample sizes are a little low here for some groups (low, high, severe) for “no” to “restricted funeral arrangements”, which is possibly why *P* = 0.018 (rather than, say, *P* < 0.001 as for social isolation and loneliness).

**Table S9: Results for subscales and scale scores for the AAG questionnaire and support needed as a function of cause of death, i.e., COVID-19 (confirmed or suspected) or non-COVID-19**

|  | | AAG | | | | Support Needed | | |
| --- | --- | --- | --- | --- | --- | --- | --- | --- |
|  | | Overwhelmed | Controlled | Reversed Resilience | IOV | Practical | Emotional | Overall |
| COVID | *n* | 306 | 305 | 305 | 303 | 302 | 304 | 304 |
|  | Mean | 8.95 | 6.40 | 5.70 | 21.07 | 2.53 | 3.55 | 3.32 |
|  | SD | 2.73 | 2.84 | 2.96 | 4.49 | 1.09 | 0.98 | 0.92 |
|  | *Q*_1_ | 7 | 4.5 | 3 | 18 | 1.67 | 3 | 2.79 |
|  | *Q*_2_ (Median) | 9 | 6 | 5 | 22 | 2.33 | 3.65 | 3.31 |
|  | *Q*_3_ | 11 | 8 | 8 | 24 | 3.33 | 4.3 | 4 |
| Non-Covid | *n* | 398 | 394 | 395 | 394 | 387 | 391 | 391 |
|  | Mean | 8.21 | 6.78 | 4.97 | 19.91 | 2.32 | 3.17 | 2.97 |
|  | SD | 2.81 | 2.6 | 2.66 | 4.93 | 1.09 | 1.13 | 1.03 |
|  | *Q*_1_ | 6 | 5 | 3 | 17 | 1.33 | 2.30 | 2.23 |
|  | *Q*_2_ (Median) | 8 | 7 | 5 | 20 | 2 | 3.2 | 3 |
|  | *Q*_3_ | 11 | 9 | 7 | 23 | 3 | 4 | 3.69 |
| Cohen’s \|*d*\| | | 0.27 | 0.14 | 0.26 | 0.25 | 0.19 | 0.36 | 0.35 |
| *t*-test: *P* = | | <0.001 | 0.068 | 0.001 | 0.001 | 0.012 | <0.001 | <0.001 |

**Table S10: Results for subscales and scale scores for the AAG questionnaire and support needed as a function of the question “Did you expect your loved one to die around this time? (e.g. if they had a terminal illness)”**

|  | | AAG | | | | Support Needed | | |
| --- | --- | --- | --- | --- | --- | --- | --- | --- |
|  | | Overwhelmed | Controlled | Reversed Resilience | IOV | Practical | Emotional | Overall |
| Yes | *n* | 113 | 113 | 113 | 113 | 108 | 110 | 110 |
|  | Mean | 7.56 | 6.91 | 4.35 | 18.81 | 2.15 | 2.98 | 2.787 |
|  | SD | 2.75 | 2.31 | 2.58 | 4.83 | 1.01 | 1.03 | 0.96 |
|  | Median | 8 | 7 | 4 | 19 | 2 | 3 | 2.82 |
| No | *n* | 547 | 543 | 543 | 541 | 538 | 541 | 541 |
|  | Mean | 8.83 | 6.5 | 5.56 | 20.88 | 2.49 | 3.45 | 3.23 |
|  | SD | 2.74 | 2.79 | 2.84 | 4.72 | 1.11 | 1.07 | 0.99 |
|  | Median | 9 | 7 | 5 | 21 | 2.33 | 3.6 | 3.25 |
| Cohen’s \|*d*\| | | 0.47 | 0.36 | 0.46 | 0.46 | 0.33 | 0.58 | 0.57 |
| *P* = | | <0.001 | 0.032 | <0.001 | <0.001 | 0.002 | <0.001 | <0.001 |

**Figure S1: Scatter plots showing the IOV and overall support score as a function of the age of the bereaved person. Quadratic line fits with associated 95% confidence intervals of the estimate have been added to these figures also in order to show the general trend of a minimum in the outcomes at about an age of 50 years old more clearly.**

IOV

| Age of the Bereaved | Data |
| --- | --- |
| 35 | 20 |
| 33 | 21 |
| 63 | 20 |
| 44 | 25 |
| 34 | 16 |
| 52 | 24 |
| 44 | 22 |
| 53 | 18 |
| 22 | 28 |
| 24 | 13 |
| 31 | 23 |
| 32 | 21 |
| 28 | 24 |
| 18 | 19 |
| 54 | 21 |
| 56 | 22 |
| 29 | 25 |
| 28 | 20 |
| 40 | 24 |
| 28 | 21 |
| 37 | 28 |
| 60 | 27 |
| 60 | 22 |
| 48 | 24 |
| 57 | 24 |
| 68 | 22 |
| 34 | 20 |
| 38 | 19 |
| 57 | 25 |
| 60 | 19 |
| 30 | 23 |
| 55 | 19 |
| 69 | 30 |
| 38 | 18 |
| 44 | 17 |
| 41 | 16 |
| 72 | 29 |
| 39 | 12 |
| 72 | 25 |
| 67 | 26 |
| 44 | 19 |
| 46 | 19 |
| 43 | 17 |
| 48 | 18 |
| 23 | 18 |
| 47 | 19 |
| 50 | 22 |
| 45 | 27 |
| 43 | 25 |
| 36 | 17 |
| 57 | 18 |
| 47 | 21 |
| 41 | 18 |
| 25 | 26 |
| 18 | 25 |
| 45 | 9 |
| 32 | 22 |
| 46 | 20 |
| 54 | 24 |
| 53 | 21 |
| 50 | 11 |
| 42 | 18 |
| 48 | 19 |
| 53 | 16 |
| 60 | 27 |
| 54 | 23 |
| 21 | 16 |
| 47 | 23 |
| 47 | 17 |
| 48 | 22 |
| 52 | 21 |
| 56 | 24 |
| 60 | 21 |
| 44 | 18 |
| 54 | 23 |
| 50 | 26 |
| 53 | 25 |
| 49 | 18 |
| 50 | 29 |
| 53 | 11 |
| 26 | 30 |
| 25 | 24 |
| 47 | 22 |
| 55 | 30 |
| 53 | 21 |
| 53 | 20 |
| 56 | 15 |
| 31 | 17 |
| 44 | 21 |
| 56 | 21 |
| 44 | 24 |
| 44 | 23 |
| 24 | 24 |
| 28 | 20 |
| 54 | 20 |
| 30 | 29 |
| 39 | 27 |
| 48 | 4 |
| 53 | 13 |
| 30 | 26 |
| 56 | 17 |
| 54 | 24 |
| 55 | 25 |
| 57 | 15 |
| 26 | 22 |
| 67 | 23 |
| 48 | 29 |
| 26 | 12 |
| 26 | 26 |
| 55 | 12 |
| 27 | 28 |
| 38 | 18 |
| 55 | 15 |
| 50 | 26 |
| 60 | 26 |
| 39 | 17 |
| 55 | 25 |
| 56 | 23 |
| 61 | 25 |
| 58 | 16 |
| 49 | 21 |
|  | 27 |
| 55 | 20 |
| 58 | 20 |
| 66 | 22 |
| 58 | 23 |
| 58 | 21 |
| 58 | 16 |
| 36 | 24 |
| 63 | 23 |
| 68 | 22 |
| 34 | 23 |
| 70 | 23 |
| 60 | 19 |
| 29 | 21 |
| 57 | 21 |
| 70 | 18 |
| 26 | 11 |
| 60 | 24 |
| 41 | 25 |
| 36 | 21 |
| 23 | 25 |
| 34 | 24 |
| 26 | 18 |
| 49 | 26 |
| 34 | 24 |
| 37 | 28 |
| 70 | 22 |
| 55 | 24 |
| 55 | 17 |
| 57 | 28 |
| 62 | 14 |
| 38 | 12 |
| 31 | 22 |
| 28 | 20 |
| 58 | 23 |
| 61 | 24 |
| 32 | 17 |
| 34 | 14 |
| 63 | 21 |
| 33 | 24 |
| 29 | 22 |
| 59 | 14 |
| 55 | 24 |
| 63 | 23 |
| 61 | 26 |
| 28 | 20 |
| 36 | 23 |
| 39 | 21 |
| 39 | 27 |
| 63 | 22 |
| 31 | 21 |
| 38 | 23 |
| 30 | 19 |
| 32 | 25 |
| 63 | 20 |
| 63 | 20 |
| 57 | 20 |
| 58 | 15 |
| 38 | 27 |
| 63 | 15 |
| 41 | 20 |
| 37 | 23 |
| 58 | 20 |
| 40 | 23 |
| 31 | 23 |
| 55 | 19 |
| 29 | 23 |
| 38 | 17 |
| 61 | 26 |
| 37 | 18 |
| 39 | 21 |
| 67 | 20 |
| 43 | 25 |
| 61 | 13 |

| Age of the Bereaved | Quadratic Line Fit | 95% CI |  |
| --- | --- | --- | --- |
| 23 | 21.642 | 20.4765 | 22.8075 |
| 23 | 21.642 | 20.4765 | 22.8075 |
| 23 | 21.642 | 20.4765 | 22.8075 |
| 24 | 21.5196 | 20.4304 | 22.6088 |
| 24 | 21.5196 | 20.4304 | 22.6088 |
| 24 | 21.5196 | 20.4304 | 22.6088 |
| 24 | 21.5196 | 20.4304 | 22.6088 |
| 24 | 21.5196 | 20.4304 | 22.6088 |
| 24 | 21.5196 | 20.4304 | 22.6088 |
| 25 | 21.402 | 20.3851 | 22.4188 |
| 25 | 21.402 | 20.3851 | 22.4188 |
| 25 | 21.402 | 20.3851 | 22.4188 |
| 26 | 21.2891 | 20.3405 | 22.2377 |
| 26 | 21.2891 | 20.3405 | 22.2377 |
| 26 | 21.2891 | 20.3405 | 22.2377 |
| 26 | 21.2891 | 20.3405 | 22.2377 |
| 26 | 21.2891 | 20.3405 | 22.2377 |
| 26 | 21.2891 | 20.3405 | 22.2377 |
| 26 | 21.2891 | 20.3405 | 22.2377 |
| 26 | 21.2891 | 20.3405 | 22.2377 |
| 26 | 21.2891 | 20.3405 | 22.2377 |
| 26 | 21.2891 | 20.3405 | 22.2377 |
| 27 | 21.1811 | 20.2966 | 22.0656 |
| 27 | 21.1811 | 20.2966 | 22.0656 |
| 27 | 21.1811 | 20.2966 | 22.0656 |
| 27 | 21.1811 | 20.2966 | 22.0656 |
| 27 | 21.1811 | 20.2966 | 22.0656 |
| 28 | 21.0779 | 20.2531 | 21.9026 |
| 28 | 21.0779 | 20.2531 | 21.9026 |
| 28 | 21.0779 | 20.2531 | 21.9026 |
| 28 | 21.0779 | 20.2531 | 21.9026 |
| 28 | 21.0779 | 20.2531 | 21.9026 |
| 28 | 21.0779 | 20.2531 | 21.9026 |
| 28 | 21.0779 | 20.2531 | 21.9026 |
| 28 | 21.0779 | 20.2531 | 21.9026 |
| 28 | 21.0779 | 20.2531 | 21.9026 |
| 28 | 21.0779 | 20.2531 | 21.9026 |
| 28 | 21.0779 | 20.2531 | 21.9026 |
| 29 | 20.9794 | 20.21 | 21.7487 |
| 29 | 20.9794 | 20.21 | 21.7487 |
| 29 | 20.9794 | 20.21 | 21.7487 |
| 29 | 20.9794 | 20.21 | 21.7487 |
| 29 | 20.9794 | 20.21 | 21.7487 |
| 29 | 20.9794 | 20.21 | 21.7487 |
| 29 | 20.9794 | 20.21 | 21.7487 |
| 29 | 20.9794 | 20.21 | 21.7487 |
| 29 | 20.9794 | 20.21 | 21.7487 |
| 29 | 20.9794 | 20.21 | 21.7487 |
| 30 | 20.8857 | 20.1672 | 21.6042 |
| 30 | 20.8857 | 20.1672 | 21.6042 |
| 30 | 20.8857 | 20.1672 | 21.6042 |
| 30 | 20.8857 | 20.1672 | 21.6042 |
| 30 | 20.8857 | 20.1672 | 21.6042 |
| 30 | 20.8857 | 20.1672 | 21.6042 |
| 30 | 20.8857 | 20.1672 | 21.6042 |
| 30 | 20.8857 | 20.1672 | 21.6042 |
| 30 | 20.8857 | 20.1672 | 21.6042 |
| 30 | 20.8857 | 20.1672 | 21.6042 |
| 30 | 20.8857 | 20.1672 | 21.6042 |
| 31 | 20.7968 | 20.1246 | 21.469 |
| 31 | 20.7968 | 20.1246 | 21.469 |
| 31 | 20.7968 | 20.1246 | 21.469 |
| 31 | 20.7968 | 20.1246 | 21.469 |
| 31 | 20.7968 | 20.1246 | 21.469 |
| 31 | 20.7968 | 20.1246 | 21.469 |
| 31 | 20.7968 | 20.1246 | 21.469 |
| 31 | 20.7968 | 20.1246 | 21.469 |
| 31 | 20.7968 | 20.1246 | 21.469 |
| 32 | 20.7127 | 20.0821 | 21.3433 |
| 32 | 20.7127 | 20.0821 | 21.3433 |
| 32 | 20.7127 | 20.0821 | 21.3433 |
| 32 | 20.7127 | 20.0821 | 21.3433 |
| 32 | 20.7127 | 20.0821 | 21.3433 |
| 32 | 20.7127 | 20.0821 | 21.3433 |
| 32 | 20.7127 | 20.0821 | 21.3433 |
| 32 | 20.7127 | 20.0821 | 21.3433 |
| 32 | 20.7127 | 20.0821 | 21.3433 |
| 32 | 20.7127 | 20.0821 | 21.3433 |
| 33 | 20.6334 | 20.0397 | 21.227 |
| 33 | 20.6334 | 20.0397 | 21.227 |
| 33 | 20.6334 | 20.0397 | 21.227 |
| 33 | 20.6334 | 20.0397 | 21.227 |
| 34 | 20.5588 | 19.9973 | 21.1203 |
| 34 | 20.5588 | 19.9973 | 21.1203 |
| 34 | 20.5588 | 19.9973 | 21.1203 |
| 34 | 20.5588 | 19.9973 | 21.1203 |
| 34 | 20.5588 | 19.9973 | 21.1203 |
| 34 | 20.5588 | 19.9973 | 21.1203 |
| 34 | 20.5588 | 19.9973 | 21.1203 |
| 34 | 20.5588 | 19.9973 | 21.1203 |
| 34 | 20.5588 | 19.9973 | 21.1203 |
| 34 | 20.5588 | 19.9973 | 21.1203 |
| 34 | 20.5588 | 19.9973 | 21.1203 |
| 35 | 20.489 | 19.955 | 21.0231 |
| 35 | 20.489 | 19.955 | 21.0231 |
| 35 | 20.489 | 19.955 | 21.0231 |
| 35 | 20.489 | 19.955 | 21.0231 |
| 35 | 20.489 | 19.955 | 21.0231 |
| 36 | 20.4241 | 19.9131 | 20.9351 |
| 36 | 20.4241 | 19.9131 | 20.9351 |
| 36 | 20.4241 | 19.9131 | 20.9351 |
| 36 | 20.4241 | 19.9131 | 20.9351 |
| 36 | 20.4241 | 19.9131 | 20.9351 |
| 36 | 20.4241 | 19.9131 | 20.9351 |
| 36 | 20.4241 | 19.9131 | 20.9351 |
| 36 | 20.4241 | 19.9131 | 20.9351 |
| 36 | 20.4241 | 19.9131 | 20.9351 |
| 36 | 20.4241 | 19.9131 | 20.9351 |
| 36 | 20.4241 | 19.9131 | 20.9351 |
| 36 | 20.4241 | 19.9131 | 20.9351 |
| 37 | 20.3639 | 19.8716 | 20.8561 |
| 37 | 20.3639 | 19.8716 | 20.8561 |
| 37 | 20.3639 | 19.8716 | 20.8561 |
| 37 | 20.3639 | 19.8716 | 20.8561 |
| 37 | 20.3639 | 19.8716 | 20.8561 |
| 37 | 20.3639 | 19.8716 | 20.8561 |
| 37 | 20.3639 | 19.8716 | 20.8561 |
| 37 | 20.3639 | 19.8716 | 20.8561 |
| 37 | 20.3639 | 19.8716 | 20.8561 |
| 37 | 20.3639 | 19.8716 | 20.8561 |
| 37 | 20.3639 | 19.8716 | 20.8561 |
| 38 | 20.3085 | 19.8311 | 20.7859 |
| 38 | 20.3085 | 19.8311 | 20.7859 |
| 38 | 20.3085 | 19.8311 | 20.7859 |
| 38 | 20.3085 | 19.8311 | 20.7859 |
| 38 | 20.3085 | 19.8311 | 20.7859 |
| 38 | 20.3085 | 19.8311 | 20.7859 |
| 38 | 20.3085 | 19.8311 | 20.7859 |
| 38 | 20.3085 | 19.8311 | 20.7859 |
| 38 | 20.3085 | 19.8311 | 20.7859 |
| 38 | 20.3085 | 19.8311 | 20.7859 |
| 38 | 20.3085 | 19.8311 | 20.7859 |
| 38 | 20.3085 | 19.8311 | 20.7859 |
| 38 | 20.3085 | 19.8311 | 20.7859 |
| 38 | 20.3085 | 19.8311 | 20.7859 |
| 38 | 20.3085 | 19.8311 | 20.7859 |
| 38 | 20.3085 | 19.8311 | 20.7859 |
| 38 | 20.3085 | 19.8311 | 20.7859 |
| 39 | 20.2579 | 19.7918 | 20.7239 |
| 39 | 20.2579 | 19.7918 | 20.7239 |
| 39 | 20.2579 | 19.7918 | 20.7239 |
| 39 | 20.2579 | 19.7918 | 20.7239 |
| 39 | 20.2579 | 19.7918 | 20.7239 |
| 39 | 20.2579 | 19.7918 | 20.7239 |
| 39 | 20.2579 | 19.7918 | 20.7239 |
| 39 | 20.2579 | 19.7918 | 20.7239 |
| 39 | 20.2579 | 19.7918 | 20.7239 |
| 39 | 20.2579 | 19.7918 | 20.7239 |
| 39 | 20.2579 | 19.7918 | 20.7239 |
| 39 | 20.2579 | 19.7918 | 20.7239 |
| 39 | 20.2579 | 19.7918 | 20.7239 |
| 39 | 20.2579 | 19.7918 | 20.7239 |
| 39 | 20.2579 | 19.7918 | 20.7239 |
| 39 | 20.2579 | 19.7918 | 20.7239 |
| 39 | 20.2579 | 19.7918 | 20.7239 |
| 40 | 20.212 | 19.7543 | 20.6698 |
| 40 | 20.212 | 19.7543 | 20.6698 |
| 40 | 20.212 | 19.7543 | 20.6698 |
| 40 | 20.212 | 19.7543 | 20.6698 |
| 40 | 20.212 | 19.7543 | 20.6698 |
| 40 | 20.212 | 19.7543 | 20.6698 |
| 40 | 20.212 | 19.7543 | 20.6698 |
| 40 | 20.212 | 19.7543 | 20.6698 |
| 40 | 20.212 | 19.7543 | 20.6698 |
| 40 | 20.212 | 19.7543 | 20.6698 |
| 40 | 20.212 | 19.7543 | 20.6698 |
| 40 | 20.212 | 19.7543 | 20.6698 |
| 40 | 20.212 | 19.7543 | 20.6698 |
| 40 | 20.212 | 19.7543 | 20.6698 |
| 41 | 20.171 | 19.719 | 20.623 |
| 41 | 20.171 | 19.719 | 20.623 |
| 41 | 20.171 | 19.719 | 20.623 |
| 41 | 20.171 | 19.719 | 20.623 |
| 41 | 20.171 | 19.719 | 20.623 |
| 41 | 20.171 | 19.719 | 20.623 |
| 41 | 20.171 | 19.719 | 20.623 |
| 41 | 20.171 | 19.719 | 20.623 |
| 41 | 20.171 | 19.719 | 20.623 |
| 41 | 20.171 | 19.719 | 20.623 |
| 41 | 20.171 | 19.719 | 20.623 |
| 41 | 20.171 | 19.719 | 20.623 |
| 41 | 20.171 | 19.719 | 20.623 |
| 41 | 20.171 | 19.719 | 20.623 |
| 41 | 20.171 | 19.719 | 20.623 |
| 41 | 20.171 | 19.719 | 20.623 |
| 41 | 20.171 | 19.719 | 20.623 |
| 41 | 20.171 | 19.719 | 20.623 |
| 41 | 20.171 | 19.719 | 20.623 |
| 42 | 20.1347 | 19.6865 | 20.5829 |
| 42 | 20.1347 | 19.6865 | 20.5829 |
| 42 | 20.1347 | 19.6865 | 20.5829 |
| 42 | 20.1347 | 19.6865 | 20.5829 |
| 42 | 20.1347 | 19.6865 | 20.5829 |
| 42 | 20.1347 | 19.6865 | 20.5829 |
| 42 | 20.1347 | 19.6865 | 20.5829 |
| 42 | 20.1347 | 19.6865 | 20.5829 |
| 42 | 20.1347 | 19.6865 | 20.5829 |
| 43 | 20.1033 | 19.6573 | 20.5492 |
| 43 | 20.1033 | 19.6573 | 20.5492 |
| 43 | 20.1033 | 19.6573 | 20.5492 |

Overall Support Needs

| Age of the Bereaved | Data |
| --- | --- |
| 35 | 3.62 |
| 33 | 5 |
| 63 | 2.31 |
| 44 | 2.92 |
| 34 | 4 |
| 52 | 4.31 |
| 44 | 2.46 |
| 53 | 4.31 |
| 22 | 3.46 |
| 24 | 1.77 |
| 31 | 3.85 |
| 32 | 2.31 |
| 28 | 3.31 |
| 18 | 4.31 |
| 54 | 3 |
| 56 | 2.92 |
| 29 | 3.46 |
| 28 | 2.92 |
| 40 | 4.69 |
| 28 | 2.92 |
| 37 | 4.25 |
| 60 | 4.69 |
| 60 | 4.69 |
| 48 | 2.77 |
| 57 | 4.45 |
| 68 | 2.38 |
| 34 | 4.46 |
| 38 | 3.85 |
| 57 | 3.92 |
| 60 | 4 |
| 30 | 3.69 |
| 55 | 3.23 |
| 69 | 3.46 |
| 38 | 2.8 |
| 44 | 2 |
| 41 | 2.62 |
| 72 | 4.91 |
| 39 | 2.69 |
| 72 | 2.54 |
| 67 | 4.69 |
| 44 | 2.54 |
| 46 | 2.92 |
| 43 | 3.31 |
| 48 | 4.31 |
| 23 | 4.54 |
| 47 | 2.85 |
| 50 | 4.31 |
| 45 | 5 |
| 43 | 3.92 |
| 36 | 3.15 |
| 57 | 1.38 |
| 47 | 2.92 |
| 41 | 3.69 |
| 25 | 3.58 |
| 18 | 4.46 |
| 45 | 2.92 |
| 32 | 3.92 |
| 46 | 3 |
| 54 | 1 |
| 53 | 3.08 |
| 50 | 3.46 |
| 42 | 2.15 |
| 48 | 4.15 |
| 53 | 4.64 |
| 60 | 4.62 |
| 56 | 4.31 |
| 54 | 5 |
| 21 | 3.23 |
| 47 | 4.08 |
| 47 | 3.69 |
| 48 | 3.85 |
| 52 | 1.85 |
| 56 | 4.46 |
| 60 | 3.92 |
| 44 | 3 |
| 54 | 3.15 |
| 50 | 2.85 |
| 53 | 1.08 |
| 49 | 1.38 |
| 50 | 4.08 |
| 53 | 2.08 |
| 26 | 3.54 |
| 25 | 4.46 |
| 47 | 4.92 |
| 55 | 4.92 |
| 53 | 2.62 |
| 53 | 3 |
| 56 | 5 |
| 31 | 2.69 |
| 44 | 3.85 |
| 56 | 3.92 |
| 44 | 3.31 |
| 44 | 3.62 |
| 24 | 4.85 |
| 28 | 3.31 |
| 54 | 2.31 |
| 30 | 4.92 |
| 39 | 3.31 |
| 48 | 2.15 |
| 53 | 1.92 |
| 30 | 4.23 |
| 56 | 3.67 |
| 54 | 2.62 |
| 55 | 3.85 |
| 57 | 2.46 |
| 26 | 3.38 |
| 67 | 3.54 |
| 48 | 2.83 |
| 26 | 3.31 |
| 26 | 4.08 |
| 55 | 1.23 |
| 27 | 4 |
| 38 | 1.92 |
| 55 | 4.23 |
| 50 | 5 |
| 60 | 3.23 |
| 39 | 3.23 |
| 55 | 2.92 |
| 56 | 3.77 |
| 61 | 5 |
| 58 | 2.08 |
| 49 | 3 |
|  | 5 |
| 55 | 2.38 |
| 58 | 2.23 |
| 66 | 3.75 |
| 58 | 4 |
| 58 | 3.46 |
| 58 | 1.46 |
| 36 | 3.62 |
| 63 | 3.08 |
| 68 | 3.38 |
| 34 | 3.38 |
| 70 | 4.64 |
| 60 | 3.1 |
| 29 | 4.31 |
| 57 | 2.85 |
| 70 | 1.15 |
| 26 | 2.54 |
| 60 | 4.38 |
| 41 | 3.75 |
| 36 | 3.46 |
| 23 | 3.38 |
| 34 | 3.62 |
| 26 | 2.85 |
| 49 | 2.8 |
| 34 | 2.92 |
| 70 | 3.92 |
| 55 | 3.69 |
| 55 | 2.69 |
| 72 | 1.54 |
| 57 | 5 |
| 62 | 3.62 |
| 38 | 3.15 |
| 31 | 4.69 |
| 28 | 3.15 |
| 58 | 3.77 |
| 61 | 1.23 |
| 32 | 3.31 |
| 34 | 1.46 |
| 63 | 3.23 |
| 33 | 3 |
| 29 | 2.31 |
| 59 | 1.38 |
| 55 | 4.62 |
| 63 | 2.62 |
| 61 | 2.92 |
| 28 | 3.85 |
| 36 | 3.92 |
| 39 | 2.54 |
| 39 | 3.23 |
| 63 | 2.31 |
| 31 | 3.69 |
| 38 | 3.54 |
| 30 | 2.46 |
| 32 | 4 |
| 63 | 1.92 |
| 62 | 3.75 |
| 63 | 5 |
| 57 | 3.31 |
| 58 | 1.54 |
| 38 | 5 |
| 63 | 1.62 |
| 41 | 2.92 |
| 37 | 3.69 |
| 58 | 3.31 |
| 40 | 2 |
| 31 | 4.5 |
| 55 | 2.69 |
| 29 | 3.77 |
| 38 | 2.15 |
| 61 | 3.69 |
| 37 | 4.54 |
| 39 | 3 |
| 67 | 3.85 |
| 43 | 3.31 |
| 61 | 3.08 |

| Age of the Bereaved | Quadratic Line Fit | 95% CI |  |
| --- | --- | --- | --- |
| 23 | 3.50567 | 3.26109 | 3.75025 |
| 23 | 3.50567 | 3.26109 | 3.75025 |
| 23 | 3.50567 | 3.26109 | 3.75025 |
| 24 | 3.47402 | 3.24544 | 3.7026 |
| 24 | 3.47402 | 3.24544 | 3.7026 |
| 24 | 3.47402 | 3.24544 | 3.7026 |
| 24 | 3.47402 | 3.24544 | 3.7026 |
| 24 | 3.47402 | 3.24544 | 3.7026 |
| 24 | 3.47402 | 3.24544 | 3.7026 |
| 25 | 3.44345 | 3.23004 | 3.65687 |
| 25 | 3.44345 | 3.23004 | 3.65687 |
| 25 | 3.44345 | 3.23004 | 3.65687 |
| 26 | 3.41397 | 3.21487 | 3.61307 |
| 26 | 3.41397 | 3.21487 | 3.61307 |
| 26 | 3.41397 | 3.21487 | 3.61307 |
| 26 | 3.41397 | 3.21487 | 3.61307 |
| 26 | 3.41397 | 3.21487 | 3.61307 |
| 26 | 3.41397 | 3.21487 | 3.61307 |
| 26 | 3.41397 | 3.21487 | 3.61307 |
| 26 | 3.41397 | 3.21487 | 3.61307 |
| 26 | 3.41397 | 3.21487 | 3.61307 |
| 26 | 3.41397 | 3.21487 | 3.61307 |
| 27 | 3.38557 | 3.19992 | 3.57122 |
| 27 | 3.38557 | 3.19992 | 3.57122 |
| 27 | 3.38557 | 3.19992 | 3.57122 |
| 27 | 3.38557 | 3.19992 | 3.57122 |
| 27 | 3.38557 | 3.19992 | 3.57122 |
| 28 | 3.35826 | 3.18516 | 3.53135 |
| 28 | 3.35826 | 3.18516 | 3.53135 |
| 28 | 3.35826 | 3.18516 | 3.53135 |
| 28 | 3.35826 | 3.18516 | 3.53135 |
| 28 | 3.35826 | 3.18516 | 3.53135 |
| 28 | 3.35826 | 3.18516 | 3.53135 |
| 28 | 3.35826 | 3.18516 | 3.53135 |
| 28 | 3.35826 | 3.18516 | 3.53135 |
| 28 | 3.35826 | 3.18516 | 3.53135 |
| 28 | 3.35826 | 3.18516 | 3.53135 |
| 28 | 3.35826 | 3.18516 | 3.53135 |
| 29 | 3.33202 | 3.17057 | 3.49348 |
| 29 | 3.33202 | 3.17057 | 3.49348 |
| 29 | 3.33202 | 3.17057 | 3.49348 |
| 29 | 3.33202 | 3.17057 | 3.49348 |
| 29 | 3.33202 | 3.17057 | 3.49348 |
| 29 | 3.33202 | 3.17057 | 3.49348 |
| 29 | 3.33202 | 3.17057 | 3.49348 |
| 29 | 3.33202 | 3.17057 | 3.49348 |
| 29 | 3.33202 | 3.17057 | 3.49348 |
| 29 | 3.33202 | 3.17057 | 3.49348 |
| 30 | 3.30688 | 3.15612 | 3.45763 |
| 30 | 3.30688 | 3.15612 | 3.45763 |
| 30 | 3.30688 | 3.15612 | 3.45763 |
| 30 | 3.30688 | 3.15612 | 3.45763 |
| 30 | 3.30688 | 3.15612 | 3.45763 |
| 30 | 3.30688 | 3.15612 | 3.45763 |
| 30 | 3.30688 | 3.15612 | 3.45763 |
| 30 | 3.30688 | 3.15612 | 3.45763 |
| 30 | 3.30688 | 3.15612 | 3.45763 |
| 30 | 3.30688 | 3.15612 | 3.45763 |
| 30 | 3.30688 | 3.15612 | 3.45763 |
| 31 | 3.28281 | 3.14179 | 3.42383 |
| 31 | 3.28281 | 3.14179 | 3.42383 |
| 31 | 3.28281 | 3.14179 | 3.42383 |
| 31 | 3.28281 | 3.14179 | 3.42383 |
| 31 | 3.28281 | 3.14179 | 3.42383 |
| 31 | 3.28281 | 3.14179 | 3.42383 |
| 31 | 3.28281 | 3.14179 | 3.42383 |
| 31 | 3.28281 | 3.14179 | 3.42383 |
| 31 | 3.28281 | 3.14179 | 3.42383 |
| 32 | 3.25983 | 3.12758 | 3.39209 |
| 32 | 3.25983 | 3.12758 | 3.39209 |
| 32 | 3.25983 | 3.12758 | 3.39209 |
| 32 | 3.25983 | 3.12758 | 3.39209 |
| 32 | 3.25983 | 3.12758 | 3.39209 |
| 32 | 3.25983 | 3.12758 | 3.39209 |
| 32 | 3.25983 | 3.12758 | 3.39209 |
| 32 | 3.25983 | 3.12758 | 3.39209 |
| 32 | 3.25983 | 3.12758 | 3.39209 |
| 32 | 3.25983 | 3.12758 | 3.39209 |
| 33 | 3.23793 | 3.11345 | 3.36242 |
| 33 | 3.23793 | 3.11345 | 3.36242 |
| 33 | 3.23793 | 3.11345 | 3.36242 |
| 33 | 3.23793 | 3.11345 | 3.36242 |
| 34 | 3.21712 | 3.09942 | 3.33482 |
| 34 | 3.21712 | 3.09942 | 3.33482 |
| 34 | 3.21712 | 3.09942 | 3.33482 |
| 34 | 3.21712 | 3.09942 | 3.33482 |
| 34 | 3.21712 | 3.09942 | 3.33482 |
| 34 | 3.21712 | 3.09942 | 3.33482 |
| 34 | 3.21712 | 3.09942 | 3.33482 |
| 34 | 3.21712 | 3.09942 | 3.33482 |
| 34 | 3.21712 | 3.09942 | 3.33482 |
| 34 | 3.21712 | 3.09942 | 3.33482 |
| 34 | 3.21712 | 3.09942 | 3.33482 |
| 35 | 3.19739 | 3.08551 | 3.30928 |
| 35 | 3.19739 | 3.08551 | 3.30928 |
| 35 | 3.19739 | 3.08551 | 3.30928 |
| 35 | 3.19739 | 3.08551 | 3.30928 |
| 35 | 3.19739 | 3.08551 | 3.30928 |
| 36 | 3.17875 | 3.07173 | 3.28576 |
| 36 | 3.17875 | 3.07173 | 3.28576 |
| 36 | 3.17875 | 3.07173 | 3.28576 |
| 36 | 3.17875 | 3.07173 | 3.28576 |
| 36 | 3.17875 | 3.07173 | 3.28576 |
| 36 | 3.17875 | 3.07173 | 3.28576 |
| 36 | 3.17875 | 3.07173 | 3.28576 |
| 36 | 3.17875 | 3.07173 | 3.28576 |
| 36 | 3.17875 | 3.07173 | 3.28576 |
| 36 | 3.17875 | 3.07173 | 3.28576 |
| 36 | 3.17875 | 3.07173 | 3.28576 |
| 36 | 3.17875 | 3.07173 | 3.28576 |
| 37 | 3.16118 | 3.05814 | 3.26423 |
| 37 | 3.16118 | 3.05814 | 3.26423 |
| 37 | 3.16118 | 3.05814 | 3.26423 |
| 37 | 3.16118 | 3.05814 | 3.26423 |
| 37 | 3.16118 | 3.05814 | 3.26423 |
| 37 | 3.16118 | 3.05814 | 3.26423 |
| 37 | 3.16118 | 3.05814 | 3.26423 |
| 37 | 3.16118 | 3.05814 | 3.26423 |
| 37 | 3.16118 | 3.05814 | 3.26423 |
| 37 | 3.16118 | 3.05814 | 3.26423 |
| 37 | 3.16118 | 3.05814 | 3.26423 |
| 38 | 3.14471 | 3.04482 | 3.2446 |
| 38 | 3.14471 | 3.04482 | 3.2446 |
| 38 | 3.14471 | 3.04482 | 3.2446 |
| 38 | 3.14471 | 3.04482 | 3.2446 |
| 38 | 3.14471 | 3.04482 | 3.2446 |
| 38 | 3.14471 | 3.04482 | 3.2446 |
| 38 | 3.14471 | 3.04482 | 3.2446 |
| 38 | 3.14471 | 3.04482 | 3.2446 |
| 38 | 3.14471 | 3.04482 | 3.2446 |
| 38 | 3.14471 | 3.04482 | 3.2446 |
| 38 | 3.14471 | 3.04482 | 3.2446 |
| 38 | 3.14471 | 3.04482 | 3.2446 |
| 38 | 3.14471 | 3.04482 | 3.2446 |
| 38 | 3.14471 | 3.04482 | 3.2446 |
| 38 | 3.14471 | 3.04482 | 3.2446 |
| 38 | 3.14471 | 3.04482 | 3.2446 |
| 38 | 3.14471 | 3.04482 | 3.2446 |
| 39 | 3.12931 | 3.03183 | 3.22679 |
| 39 | 3.12931 | 3.03183 | 3.22679 |
| 39 | 3.12931 | 3.03183 | 3.22679 |
| 39 | 3.12931 | 3.03183 | 3.22679 |
| 39 | 3.12931 | 3.03183 | 3.22679 |
| 39 | 3.12931 | 3.03183 | 3.22679 |
| 39 | 3.12931 | 3.03183 | 3.22679 |
| 39 | 3.12931 | 3.03183 | 3.22679 |
| 39 | 3.12931 | 3.03183 | 3.22679 |
| 39 | 3.12931 | 3.03183 | 3.22679 |
| 39 | 3.12931 | 3.03183 | 3.22679 |
| 39 | 3.12931 | 3.03183 | 3.22679 |
| 39 | 3.12931 | 3.03183 | 3.22679 |
| 39 | 3.12931 | 3.03183 | 3.22679 |
| 39 | 3.12931 | 3.03183 | 3.22679 |
| 39 | 3.12931 | 3.03183 | 3.22679 |
| 39 | 3.12931 | 3.03183 | 3.22679 |
| 40 | 3.115 | 3.01929 | 3.21071 |
| 40 | 3.115 | 3.01929 | 3.21071 |
| 40 | 3.115 | 3.01929 | 3.21071 |
| 40 | 3.115 | 3.01929 | 3.21071 |
| 40 | 3.115 | 3.01929 | 3.21071 |
| 40 | 3.115 | 3.01929 | 3.21071 |
| 40 | 3.115 | 3.01929 | 3.21071 |
| 40 | 3.115 | 3.01929 | 3.21071 |
| 40 | 3.115 | 3.01929 | 3.21071 |
| 40 | 3.115 | 3.01929 | 3.21071 |
| 40 | 3.115 | 3.01929 | 3.21071 |
| 40 | 3.115 | 3.01929 | 3.21071 |
| 40 | 3.115 | 3.01929 | 3.21071 |
| 40 | 3.115 | 3.01929 | 3.21071 |
| 41 | 3.10177 | 3.0073 | 3.19624 |
| 41 | 3.10177 | 3.0073 | 3.19624 |
| 41 | 3.10177 | 3.0073 | 3.19624 |
| 41 | 3.10177 | 3.0073 | 3.19624 |
| 41 | 3.10177 | 3.0073 | 3.19624 |
| 41 | 3.10177 | 3.0073 | 3.19624 |
| 41 | 3.10177 | 3.0073 | 3.19624 |
| 41 | 3.10177 | 3.0073 | 3.19624 |
| 41 | 3.10177 | 3.0073 | 3.19624 |
| 41 | 3.10177 | 3.0073 | 3.19624 |
| 41 | 3.10177 | 3.0073 | 3.19624 |
| 41 | 3.10177 | 3.0073 | 3.19624 |
| 41 | 3.10177 | 3.0073 | 3.19624 |
| 41 | 3.10177 | 3.0073 | 3.19624 |
| 41 | 3.10177 | 3.0073 | 3.19624 |
| 41 | 3.10177 | 3.0073 | 3.19624 |
| 41 | 3.10177 | 3.0073 | 3.19624 |
| 41 | 3.10177 | 3.0073 | 3.19624 |
| 41 | 3.10177 | 3.0073 | 3.19624 |
| 42 | 3.08963 | 2.99597 | 3.18329 |
| 42 | 3.08963 | 2.99597 | 3.18329 |
| 42 | 3.08963 | 2.99597 | 3.18329 |
| 42 | 3.08963 | 2.99597 | 3.18329 |
| 42 | 3.08963 | 2.99597 | 3.18329 |
| 42 | 3.08963 | 2.99597 | 3.18329 |
| 42 | 3.08963 | 2.99597 | 3.18329 |
| 42 | 3.08963 | 2.99597 | 3.18329 |
| 42 | 3.08963 | 2.99597 | 3.18329 |
| 43 | 3.07857 | 2.98539 | 3.17175 |
| 43 | 3.07857 | 2.98539 | 3.17175 |
| 43 | 3.07857 | 2.98539 | 3.17175 |
