## Supplementary material for "Factors associated with higher levels of grief and support needs among people bereaved during the pandemic: Results from a national online survey": CHERRIES reporting checklist

Table 1. Checklist for Reporting Results of Internet E-Surveys (CHERRIES)

Eysenbach G. Improving the quality of Web surveys: the Checklist for Reporting Results of Internet E-Surveys (CHERRIES). Journal of medical Internet research. 2004;6(3):e34.

|  | **Checklist for Reporting Results of Internet E-Surveys (CHERRIES)** | |
| --- | --- | --- |
| ***Item Category*** | ***Checklist Item*** | ***Explanation*** |
| **Design** |  |  |
|  | Describe survey design | Describe target population, sample frame. Is the sample a convenience sample? (In “open” surveys this is most likely.)  Page 4, Study design |
| **IRB (Institutional Review Board) approval and informed consent process** |  |  |
|  | IRB approval | Mention whether the study has been approved by an IRB.  Page 7 ethics statement |
|  | Informed consent | Describe the informed consent process. Where were the participants told the length of time of the survey, which data were stored and where and for how long, who the investigator was, and the purpose of the study?  Page 5 Study Procedure, Supplementary File 1 |
|  | Data protection | If any personal information was collected or stored, describe what mechanisms were used to protect unauthorized access.  Page 5 Study Procedure, Supplementary File 1 |
| **Development and pre-testing** |  |  |
|  | Development and testing | State how the survey was developed, including whether the usability and technical functionality of the electronic questionnaire had been tested before fielding the questionnaire.  Page 4, Survey development |
| **Recruitment process and description of the sample having access to the questionnaire** |  |  |
|  | Open survey versus closed survey | An “open survey” is a survey open for each visitor of a site, while a closed survey is only open to a sample which the investigator knows (password-protected survey).  Page 5, Study design/ Survey development-open survey |
|  | Contact mode | Indicate whether or not the initial contact with the potential participants was made on the Internet. (Investigators may also send out questionnaires by mail and allow for Web-based data entry.)  Page 5, Study Procedure-mixed contact methods |
|  | Advertising the survey | How/where was the survey announced or advertised? Some examples are offline media (newspapers), or online (mailing lists – If yes, which ones?) or banner ads (Where were these banner ads posted and what did they look like?). It is important to know the wording of the announcement as it will heavily influence who chooses to participate. Ideally the survey announcement should be published as an appendix.  Page 5-6, Study Procedure, Supplementary File 2 (advert), multiple methods used |
| **Survey administration** |  |  |
|  | Web/E-mail | State the type of e-survey (eg, one posted on a Web site, or one sent out through e-mail). If it is an e-mail survey, were the responses entered manually into a database, or was there an automatic method for capturing responses?  Page 5-6, Study Procedure, posted on website and sent out via e-mail |
|  | Context | Describe the Web site (for mailing list/newsgroup) in which the survey was posted. What is the Web site about, who is visiting it, what are visitors normally looking for? Discuss to what degree the content of the Web site could pre-select the sample or influence the results. For example, a survey about vaccination on a anti-immunization Web site will have different results from a Web survey conducted on a government Web site  Page 6 Study Procedure, website set up to host survey and other study material |
|  | Mandatory/voluntary | Was it a mandatory survey to be filled in by every visitor who wanted to enter the Web site, or was it a voluntary survey?  Page 5 Study Procedure- voluntary |
|  | Incentives | Were any incentives offered (eg, monetary, prizes, or non-monetary incentives such as an offer to provide the survey results)?  Page 5, Study Procedure- no incentives |
|  | Time/Date | In what timeframe were the data collected?  Page 5, Study Procedure |
|  | Randomization of items or questionnaires | To prevent biases items can be randomized or alternated.  Page 4, Survey Development |
|  | Adaptive questioning | Use adaptive questioning (certain items, or only conditionally displayed based on responses to other items) to reduce number and complexity of the questions.  Supplementary file 1, detailed on survey copy |
|  | Number of Items | What was the number of questionnaire items per page? The number of items is an important factor for the completion rate.  Supplementary file 1, detailed on survey copy |
|  | Number of screens (pages) | Over how many pages was the questionnaire distributed? The number of items is an important factor for the completion rate.  Supplementary file 1, detailed on survey copy |
|  | Completeness check | It is technically possible to do consistency or completeness checks before the questionnaire is submitted. Was this done, and if “yes”, how (usually JAVAScript)? An alternative is to check for completeness after the questionnaire has been submitted (and highlight mandatory items). If this has been done, it should be reported. All items should provide a non-response option such as “not applicable” or “rather not say”, and selection of one response option should be enforced.  Supplementary file 1, Survey- explains all questions apart for consent are optional. |
|  | Review step | State whether respondents were able to review and change their answers (eg, through a Back button or a Review step which displays a summary of the responses and asks the respondents if they are correct).  Supplementary file 1, Instructions for completion at front of survey |
| **Response rates** |  |  |
|  | Unique site visitor | If you provide view rates or participation rates, you need to define how you determined a unique visitor. There are different techniques available, based on IP addresses or cookies or both.  N/A-Jisc survey doesn’t provide this information |
|  | View rate (Ratio of unique survey visitors/unique site visitors) | Requires counting unique visitors to the first page of the survey, divided by the number of unique site visitors (not page views!). It is not unusual to have view rates of less than 0.1 % if the survey is voluntary.  N/A-Jisc survey doesn’t provide this information |
|  | Participation rate (Ratio of unique visitors who agreed to participate/unique first survey page visitors) | Count the unique number of people who filled in the first survey page (or agreed to participate, for example by checking a checkbox), divided by visitors who visit the first page of the survey (or the informed consents page, if present). This can also be called “recruitment” rate.  N/A-Jisc survey doesn’t provide this information |
|  | Completion rate (Ratio of users who finished the survey/users who agreed to participate) | The number of people submitting the last questionnaire page, divided by the number of people who agreed to participate (or submitted the first survey page). This is only  relevant if there is a separate “informed consent” page or if the survey goes over several pages. This is a measure for attrition. Note that “completion” can involve leaving questionnaire items blank. This is not a measure for how completely questionnaires were filled in. (If you need a measure for this, use the word “completeness rate”.)  N/A-Jisc survey doesn’t provide this information |
| **Preventing multiple entries from the same individual** |  |  |
|  | Cookies used | Indicate whether cookies were used to assign a unique user identifier to each client computer. If so, mention the page on which the cookie was set and read, and how long the cookie was valid. Were duplicate entries avoided by preventing users access to the survey twice; or were duplicate database entries having the same user ID eliminated before analysis? In the latter case, which entries were kept for analysis (eg, the first entry or the most recent)?  P6 Study Procedure, duplicates eliminated before analysis |
|  | IP check | Indicate whether the IP address of the client computer was used to identify potential duplicate entries from the same user. If so, mention the period of time for which no two entries from the same IP address were allowed (eg, 24 hours). Were duplicate entries avoided by preventing users with the same IP address access to the survey twice; or were duplicate database entries having the same IP address within a given period of time eliminated before analysis? If the latter, which entries were kept for analysis (eg, the first entry or the most recent)?  N/A -see above comment |
|  | Log file analysis | Indicate whether other techniques to analyze the log file for identification of multiple entries were used. If so, please describe.  P6 Study Procedure |
|  | Registration | In “closed” (non-open) surveys, users need to login first and it is easier to prevent duplicate entries from the same user. Describe how this was done. For example, was the survey never displayed a second time once the user had filled it in, or was the username stored together with the survey results and later eliminated? If the latter, which entries were kept for analysis (eg, the first entry or the most recent)?  N/A open survey |
| **Analysis** |  |  |
|  | Handling of incomplete questionnaires | Were only completed questionnaires analyzed? Were questionnaires which terminated early (where, for example, users did not go through all questionnaire pages) also analyzed?  P6 Study Procedure- 2 eliminated because only consent question answered |
|  | Questionnaires submitted with an atypical timestamp | Some investigators may measure the time people needed to fill in a questionnaire and exclude questionnaires that were submitted too soon. Specify the timeframe that was used as a cut-off point, and describe how this point was determined.  N/A- no time-frame used |
|  | Statistical correction | Indicate whether any methods such as weighting of items or propensity scores have been used to adjust for the non-representative sample; if so, please describe the methods.  P6 Data analysis-no correction used |
